# Imaging cortical multiple sclerosis lesions with ultra-high field MRI

**DOI:** 10.1101/2021.06.25.21259363

**Authors:** Mads A.J. Madsen, Vanessa Wiggermann, Stephan Bramow, Jeppe Romme Christensen, Finn Sellebjerg, Hartwig R. Siebner

## Abstract

**Background:** Cortical lesions are abundant in multiple sclerosis (MS), yet difficult to visualize *in vivo*. Ultra-high field (UHF) MRI at 7T and above provides technological advances suited to optimize the detection of cortical lesions in MS.

**Purpose:** To provide a narrative and quantitative systematic review of the literature on UHF MRI of cortical lesions in MS.

**Methods:** A systematic search of all literature on UHF MRI of cortical lesions in MS published before September 2020. Quantitative outcome measures included cortical lesion numbers reported using 3T and 7T MRI and between 7T MRI sequences, along with sensitivity of UHF MRI towards cortical lesions verified by histopathology.

**Results:** 7T MRI detected on average 52±26% (mean ± 95% confidence interval) more cortical lesions than the best performing image contrast at 3T, with the largest increase in type II-IV intracortical lesion detection. Across all studies, the mean cortical lesion number was 17±6/patient. In progressive MS cohorts, approximately four times more cortical lesions were reported than in CIS/early RRMS, and RRMS. Superiority of one MRI sequence over another could not be established from available data. *Post-mortem* lesion detection with UHF MRI agreed only modestly with pathological examinations. Mean pro- and retrospective sensitivity was 33±6% and 71±10%, respectively, with the highest sensitivity towards type I and type IV lesions.

**Conclusion:** UHF MRI improves cortical lesion detection in MS considerably compared to 3T MRI, particularly for type II-IV lesions. Despite modest sensitivity, 7T MRI is still capable of visualizing all aspects of cortical lesion pathology and could potentially aid clinicians in diagnosing and monitoring MS, and progressive MS in particular. However, standardization of acquisition and segmentation protocols is needed.

## 1. Introduction

Multiple sclerosis (MS) is a heterogeneous, chronic, immune-mediated disease of the central nervous system (CNS). It is characterized by widespread inflammation, demyelination and axonal degeneration causing both focal lesions and diffuse tissue alterations (Kutzelnigg et al. 2005). Magnetic resonance imaging (MRI) at field strengths up to 3T has become indispensable in the clinical management of MS, playing a key role in diagnosis, monitoring disease course and treatment response. Although conventional, clinical MRI is sensitive to white matter lesions, the diagnostic value is limited by various differential diagnoses that may underlie myelin injury in the white matter. Additionally, white matter lesions correlate relatively poorly with clinical disability (Barkhof 1999). In contrast, recent studies highlight subpial demyelination as specific for MS and rare encephalitic syndromes associated with anti-MOG-IgG (MOG-AD) (Moll et al. 2008; Popescu et al. 2010; Fischer et al. 2013; Hoftberger et al. 2020; Takai et al. 2020; Junker et al. 2020). Cortical lesions have been associated with clinical disability, lower age at death (Howell et al. 2011) and earlier conversion to secondary progressive (SP)MS (Scalfari et al. 2018; Pisani et al. 2021). Consequently, cortical lesions are now included in the 2017 revised McDonald criteria (Thompson et al. 2018). However, lesions within the cortical grey matter are difficult to identify with conventional, clinical MRI (Kidd et al. 1999; Geurts, Bo, et al. 2005; Seewann et al. 2011). A number of advances have improved cortical lesion detection at 3T, but MRI at ultra-high field (UHF, 7T and above) provides higher signal to noise ratio, allowing for sub-millimeter structural imaging (de Graaf et al. 2013). As the number of UHF MRI scanners increases worldwide, clinical applicability of these machines is increasing. In MS, numerous studies have now assessed the potential of UHF MRI for improving *in vivo* visualization of cortical lesions.

In this review, we provide a brief narrative overview of recent views on cortical pathology in MS, and the current status of cortical lesion detection at 3T. Thereafter, a systematic overview of the existing literature on imaging of cortical MS lesions with UHF MRI is presented. Specifically, we delineate the extent and distribution of cortical lesions detected with UHF MRI, highlight the utility of different MRI sequences used for lesion detection, and address the sensitivity of 7T MRI compared to histology. Finally, we discuss various approaches of cortical lesion identification and segmentation, and the potential clinical benefit of UHF MRI for cortical lesion detection.

### 1.1 Cortical involvement in MS

Pathologically, white matter plaques, i.e. confluent and sharply demarcated demyelinated areas with relative axonal sparing (Lassmann 2005), were initially identified as the hallmark of MS. However, cortical demyelination in MS was also already demonstrated more than a century ago (Hulst and Geurts 2011). In 1962, Brownell and Hughes (1962) showed in a macroscopic study that 21% of lesions were located either in the cortex or at the cortical-white matter boundary. More recent histopathologic studies demonstrated that 69% of the cortical forebrain can be demyelinated and that in primary progressive (PP)MS, the fraction of demyelinated cortex can exceed that of demyelinated white matter (Kutzelnigg et al. 2005).

Already in 1916, Dawson (1916) noted heterogeneity of cortical lesions, but it took until 1999 before staining sensitivity allowed a detailed account of the topography of cortical plaques. Cortical lesions were classified into seven subtypes depending on their spatial relationship to superficial and principal cortical veins (Kidd et al. 1999). Later, the stratification of cortical lesion types was adjusted and lesions were grouped into three currently accepted types: type I lesions are leukocortical, affecting juxta- cortical U-fibers and the deeper layers of the cortex; type II lesions are entirely intracortical; type III lesions are ribbon-shaped subpial lesions extending into the cortex in variable depths without reaching the cortical/white matter boundary (Peterson et al. 2001). A trans-cortical lesion type (type IV) spanning the entire cortex has also been described (Bo et al. 2003a; Trapp et al. 2018). However, this lesion type may be seen as a variant of type III lesions. Figure 1 shows the different lesion types as they can appear on different 7T MRI sequences. All three cortical plaque types can be found already at RRMS onset, with type I being the most frequent at this stage (Lucchinetti et al. 2011). Histopathological studies from long standing disease samples suggest that the majority of lesions are subpial (type III/IV), about 55%. (Bo et al. 2003a; Pitt et al. 2010; Peterson et al. 2001; Kilsdonk et al. 2016; Jonkman et al. 2015; Geurts, Bo, et al. 2005; Seewann et al. 2011; Schmierer, Parkes, et al. 2010). Type II intracortical lesions amount to roughly 20% and type I, i.e. leukocortical lesions, to about 25%.

**Figure 1.**
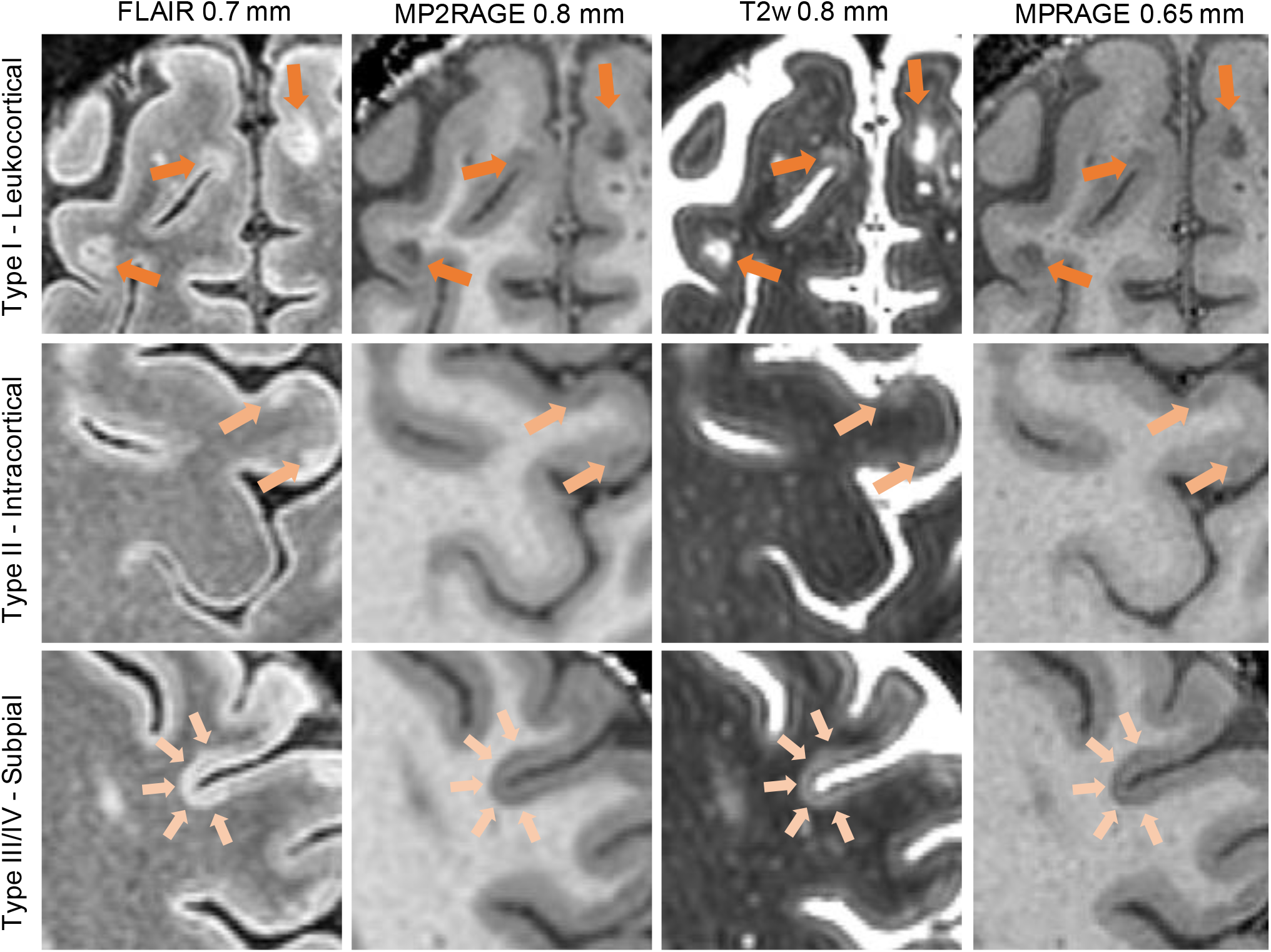
Examples of different cortical lesion types depicted on different 7T MRI sequences. Images are from a single relapsing remitting multiple sclerosis patient (male, 42 years, EDSS = 1.5, disease duration = 14 years). Images were acquired in-house and are unpublished. Abbreviations: fluid attenuated inversion recovery, MPRAGE: magnetization prepared rapid gradient echo, T2w: T2- weighted.

#### 1.1.1 Recent views on the possible pathogenesis of cortical plaques

Early histological studies on cortical lesions found that they contained fewer lymphocytes, had a less permeable blood brain barrier and showed only narrow rims of activated microglia compared to white matter lesions (Peterson et al. 2001; Bo et al. 2003b). In particular, perivascular infiltrates were uncommon and CD3-positive lymphocyte and CD68- positive microglia/macrophage densities were only a fraction of respective cell densities in white matter lesions (Peterson et al. 2001). Additionally, the presence of transected neurites and reduced neuronal density (Vercellino et al. 2005) led to the notion that neurodegenerative mechanisms might contribute independently to the formation of cortical lesions (Peterson et al. 2001; Bo et al. 2003b; Trapp et al. 2018). Yet, several lines of recent research challenge this notion: (i) It was shown that meningeal follicle-like lymphocytic infiltrates coincide with subpial cortical lesion formation (Choi et al. 2012; Howell et al. 2011; Magliozzi et al. 2007). (ii) Complement markers of classic, alternative and membrane attack complex activation are increased in cortical plaques and associated with neuronal injury (Watkins et al. 2016). (iii) Biopsies taken from early RRMS patients show signs of active demyelination in ∼ 65% of type I, ∼ 25% of type II and ∼ 20% of type III cortical plaques (Lucchinetti et al. 2011). (iv) Contrast enhancing type I lesions are found in early RRMS patients, suggestive of acute blood brain barrier disruption and a peripherally driven inflammatory response (Maranzano et al. 2017). (v) Finally, a cortical model of experimental autoimmune encephalomyelitis (EAE) described rapid type III-like subpial plaque formation associated with intracortical perivascular lymphocytic infiltrates and complement deposition (Merkler et al. 2006). Taken together, these data are consistent with an adaptive immune response driving inflammatory demyelination in both the cortex and white matter. The factors inducing cortical myelin injury and phagocytosis remain unresolved but could be related to the lymphocytic immune response surrounding the meningeal venules, which drain the cortex (Kidd et al. 1999).

### 1.2 MRI of cortical lesions at clinical field strength

Despite the detailed insights gained from histopathology, the *in vivo* implications of cortical lesions are difficult to evaluate with current clinical MRI techniques. Consequently, many efforts have been undertaken to improve cortical lesion detection with MRI. Double inversion recovery (DIR) utilizes two inversion pulses to cancel signal from both cerebrospinal fluid (CSF) and white matter (Geurts, Pouwels, et al. 2005). By cancellation of the white matter signal, DIR improves the detection of cortical lesions compared to fluid attenuated inversion recovery (FLAIR) (Geurts et al. 2011). However, DIR images suffer from a low signal to noise ratio and their specificity can be compromised by flow artifacts, among other limitations (Nelson et al. 2007; Geurts et al. 2011; Saranathan et al. 2017). Phase sensitive inversion recovery (PSIR) imaging uses phase-sensitive reconstruction along with a T1- weighted sequence to increase the range of signal intensities, which seems to improve detection of cortical lesions at 3T compared to DIR imaging (Sethi et al. 2013; Sethi et al. 2012). However, superiority of PSIR over DIR was not confirmed in recent *post-mortem* work using immunohistochemical stains (Bouman et al. 2020). Very recently, a susceptibility-weighted image contrast that utilizes signal inversion (IR-SWIET) has been proposed as another promising avenue for cortical lesion visualization at 3T (Beck et al. 2020).

Despite these efforts, prospective sensitivity remains poor at 3T (18-23% for DIR in *post-mortem* studies including histological validation (Seewann et al. 2012; Bouman et al. 2020)). Using multiple sequences for lesion segmentation increases lesion detection rates (Nelson et al. 2008; Maranzano et al. 2016), but further improvements for the *in vivo* depiction of cortical lesions are still warranted in order to accelerate correct diagnosis and management of the MS spectrum.

## 2. Methods

### 2.1 Systematic literature search of cortical lesions imaged at ultra-high field

To retrieve all relevant publications, we combined the search terms “Multiple Sclerosis” OR “MS” AND “7T” OR “7 T” OR “7Tesla” OR “7 Tesla” OR “7.0-T*” OR “ultra-high field” AND “cortical” AND “lesion*” in PubMed. The time-period covered in the search included all peer-reviewed publications up until September 1^st^, 2020. Identification, screening, eligibility and inclusion procedures followed the Preferred Reporting Items for Systematic Reviews and Meta-analyses (PRISMA) 2009 flow diagram (Moher et al. 2009). Two readers (MAJM and VW) independently screened the abstracts of all identified publications. Studies that fulfilled the following criteria were included in the qualitative analysis: 1) peer-reviewed, original articles written in English; 2) a study population including human patients diagnosed with MS; 3) investigation of cortical lesions using structural, UHF MRI. Review articles and commentaries were excluded prior to full-text assessment.

For the quantitative analysis, included manuscripts were assessed with regards to four points of interest: i) comparison of cortical lesion counts between 7T and 3T MRI, both *in vivo* and *post- mortem*; ii) *in vivo* MRI studies of adult-onset MS patients reporting total, mean or median and range counts of cortical lesions from whole- or supratentorial brain scans; iii) comparisons of cortical lesion counts between one or more 7T MRI sequences both *in vivo* and *post-mortem* and iv) the sensitivity of *post-mortem* cortical lesion detection by UHF MRI, before and after knowledge of subsequent histopathology of the same tissue blocks. Duplicate data were identified either in text or through contact with the corresponding author(s) and included based on whether studies reported mean cortical lesion count, and secondarily based on the largest cohort. Any disagreement between the two readers was followed up by HRS.

### 2.2 Outcome variables

Data extraction was done by MAJM under supervision of VW and HRS. Outcome variables included cortical lesion count for the entire MS population and for the separate MS phenotypes, where available. Due to a low study sample size, MS phenotypes were grouped into three groups: *early MS (eRRMS)*, including patients with clinically isolated syndrome (CIS) and RRMS patients < 5 years from diagnosis, *RRMS* ≥ 5 years from diagnosis, and *progressive MS (PMS)*, i.e. including both SPMS and PPMS patients. Additionally, lesion counts for individual cortical lesion types were extracted and lesion type ratios calculated, where applicable. Studies often collapsed type III and IV into a subpial category, or used a cruder classification pattern of intracortical (type II-IV) and leukocortical (type I) lesions (Treaba et al. 2019). Thus, to allow for between study comparisons, we analyzed data using both methods.

If a study only reported pooled total cortical lesion counts from its entire MS population, the group mean was calculated by dividing the count by the number of patients. If only the median and range of cortical lesion counts were reported, the group mean was estimated using the method by (Luo et al. 2018), which uses the median and range together with the sample size to estimate the sample mean. Differences in field strength performance were computed as the ratio of total cortical lesion counts between each reported sequence at 7T and the most sensitive 3T sequence within each study. This analysis was also done for lesion subtypes, where available. Sequence performance at 7T was evaluated by computing ratios of mean total cortical lesion counts and lesion subtype ratios relative to the best performing sequence within each study. For histological examinations, retrospective and/or prospective sensitivity of cortical lesion counts, and lesion type ratios were extracted.

## 3. Results

Figure 2 depicts the PRISMA flow chart and screening procedures for study inclusion. A total of 144 articles were screened by title and abstract. Of these, 84 articles, which did not fulfill our inclusion criteria, were excluded. Another six publications were excluded after full-text screening because they did not investigate cortical lesions. The remaining 54 articles were included in the qualitative analysis. For quantitative data extraction, 20 of these 54 studies were excluded for reasons described in figure 2. 34 articles remained for the quantitative analysis. Of these, 25 were included in the *in vivo* lesion count analysis. Eight articles were included in the field strength comparison (one *post-mortem*, seven *in vivo*), eight publications in the sequence comparison (three *post-mortem*, five *in vivo*) and five articles that validated UHF MRI data against histology. Among the 25 *in vivo* studies, sample size ranged from 8- 90 patients, mean age from 36-54 years and female proportion from 33-81%. Median expanded disability status scale (EDSS) was between 1.5-5 and mean disease duration ranged from 2.5-24 years. All *in vivo* studies were carried out at 7T.

**Figure 2:**
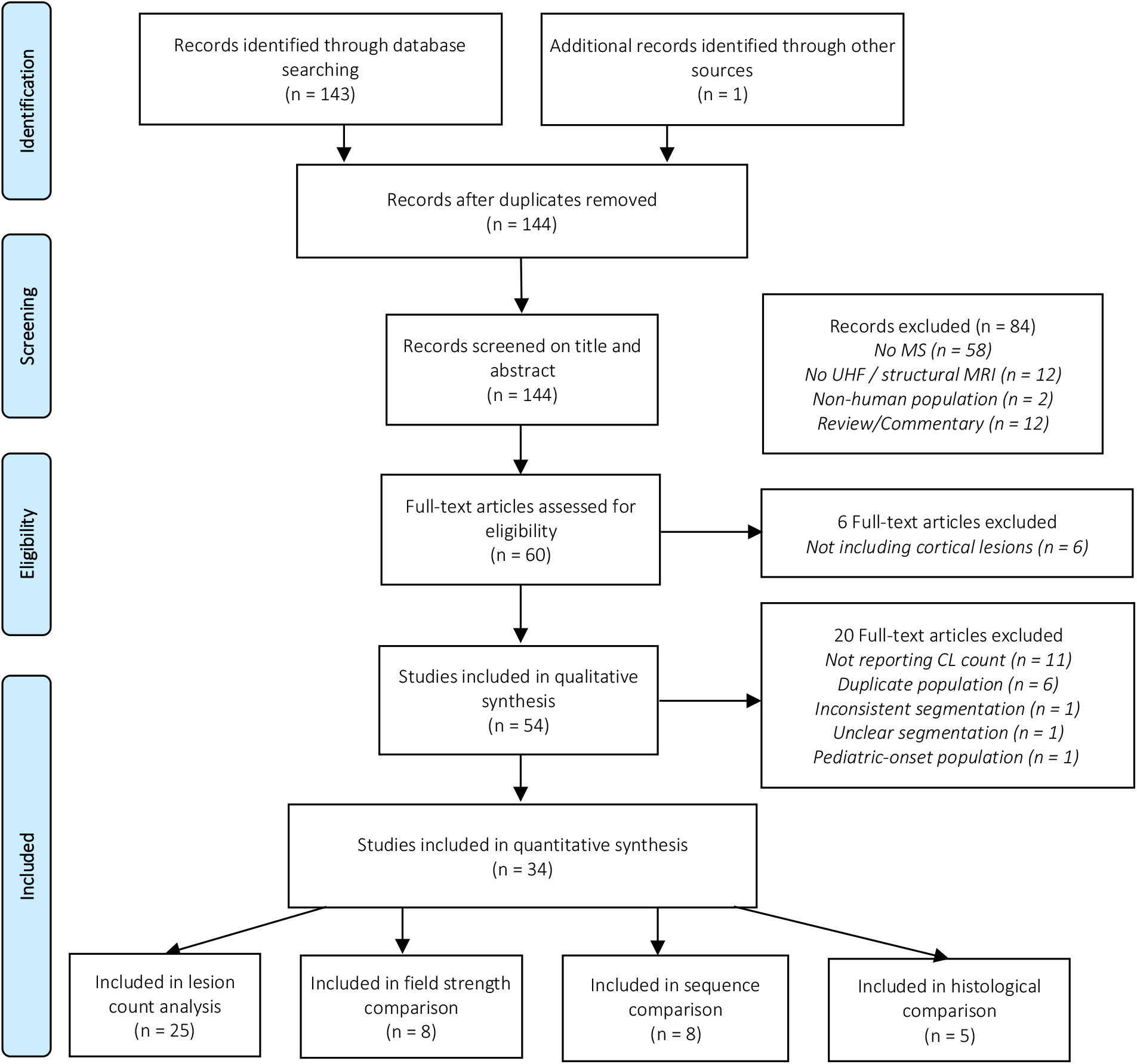
Prisma reporting chart. Initial screening excluded all reviews and commentaries and all publications that did not include MS, structural MRI at 7T or above, or included only non-human populations. Further articles were excluded if they did not assess cortical lesions. All remaining studies were included in the qualitative synthesis. Further quantitative analysis was performed by four synthesis points. Note that publications may be included in more than one of the quantitative synthesis points, shown in the bottom row.

### 3.1 Detection and segmentation of cortical lesions at 7T

The MRI sequences and contrasts used for detection and segmentation of cortical lesions varied considerably across studies. Most commonly, T2*weighted (w) images were used (n = 17), followed by T1w images (n = 7), T2w images (n = 6) or multiple sequences together (n = 9). Only one study investigated the potential of 7T MRI for automated cortical lesion detection (Fartaria et al. 2019). With a sensitivity of 58% and a false-positive rate of 40%, automated segmentation of cortical lesions was still far from a performance level that would be clinically acceptable. In all other studies, cortical lesions were manually segmented. The specific segmentation procedure was only disclosed in 32 of 54 studies. Of these, the majority of studies used two or more trained readers (n = 16), who scored lesions by consensus or segmented images independently, reaching consensus thereafter (n = 13). 11 studies did not report the number of readers.

Cortical lesions were mostly defined as being clearly demarked on the MR image and to span at least three adjacent voxels in plane, in either one (n = 8) or two consecutive slices (n = 11). Five studies used a contrast threshold of 15% compared to adjacent grey matter. Seven studies reported that hyper-/hypointensities should not be classified as cortical lesions if they appeared linear or tubular, to avoid segmentation of blood vessels (Fartaria et al. 2019; Datta et al. 2017). Five studies used the DIR consensus guidelines that were proposed for 1.5 and 3T MRI (Geurts et al. 2011). Segmentation was most often carried out in a designated MRI viewer (e.g. MIPAV, ImageJ, Slicer, Display). In two studies, it was proposed that images should be viewed in the axial plane and that lesions should be visible in the two orthogonal planes.

### 3.2 Comparison of cortical lesion detection at 7T with clinical field strengths

Eight studies performed a quantitative analysis to compare detection performance of 7T and 3T MRI, using various image contrasts. The 7T sequences detected on average 52% more cortical lesions (95% CI [26-77%]) than the best performing 3T sequence. Considering only the best performing 7T sequence for each study, cortical lesion detection improved on average by 73% (95% CI [30- 117%]) compared to the best performing 3T method (figure 3). DIR was the best performing 3T sequence in four studies, using multiple image contrasts in two studies, and T2w and magnetization prepared 2 rapid gradient echo (MP2RAGE) each performed best in one study (figure 3).

**Figure 3:**
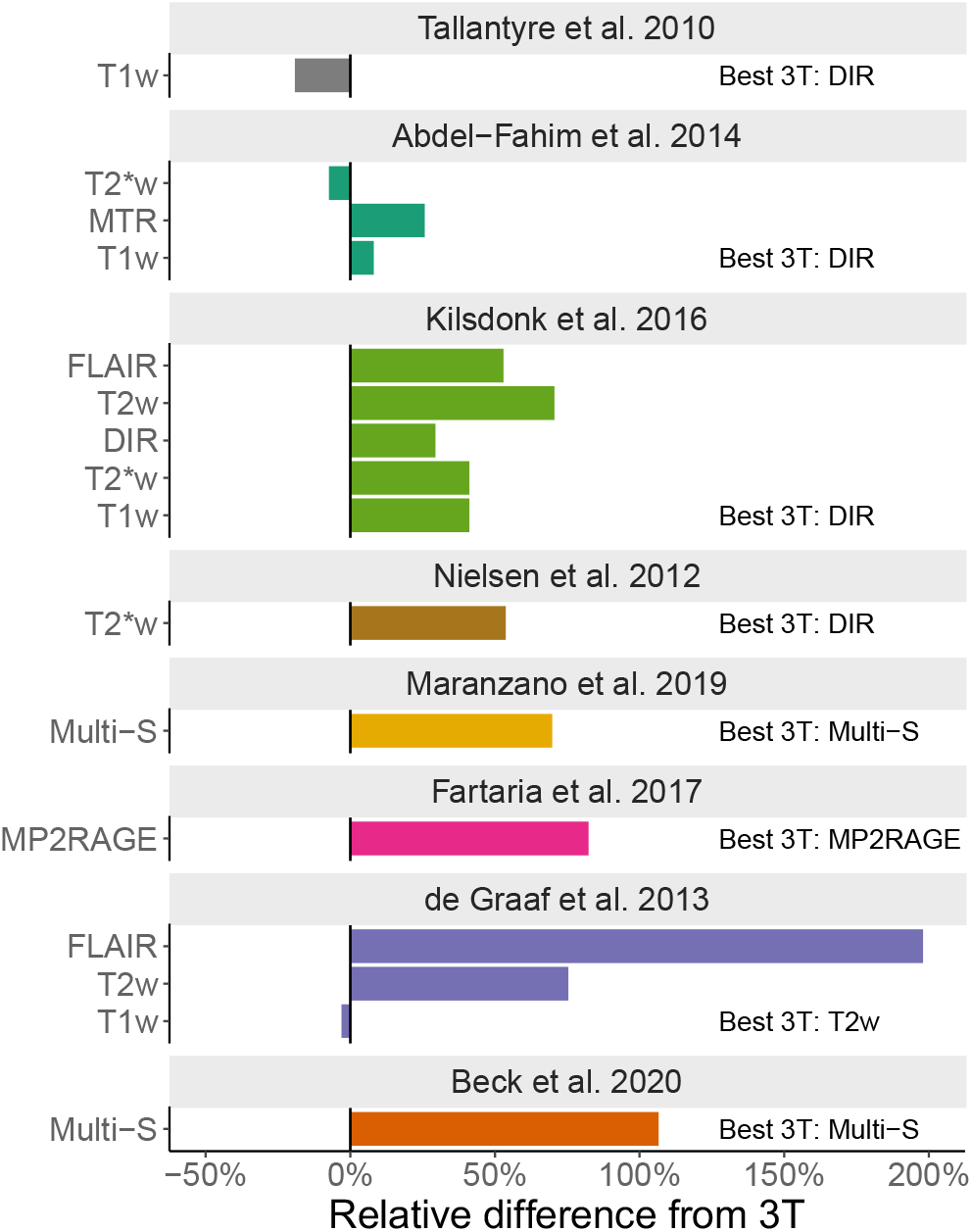
Overview of studies directly comparing cortical lesion detection counts between 3T and 7T MRI. Data shown reflect the percent difference in lesion counts relative to the 3T sequence that detected the highest number of cortical lesions within each study. Abbreviations: T1w = T1-weighted, T2*w = T2*-weighted, MTR = Magnetization Transfer Ratio, FLAIR = Fluid attenuated inversion recovery, T2w = T2-weighted, DIR = Double inversion recovery, Multi-S = Multiple sequences.

All studies reported cortical lesion subtype counts from both 3T and 7T. The increase in lesion detection at 7T MRI was particularly prominent for type II-IV intracortical lesions (166% increase in detection performance, 95% CI [39-294%]), whereas the increase in detection rate was modest for leukocortical type I lesions with an average increase in cortical lesion detection of 26% (95% CI [-13- 65%]).

### 3.3 Extent and distribution of cortical lesions detected with 7T MRI

Three studies investigated the topographical cortical lesion distribution with 7T MRI. In line with *post-mortem* histopathological data (Kutzelnigg et al. 2005; Haider et al. 2016), these studies described a predilection for sulci and deep parts of the gyri as opposed to more superficial parts of the gyri close to the hemispheric surface. Only a slight regional preponderance in temporal, frontal motor and parietal sensory areas was reported (Treaba et al. 2019; Mainero et al. 2009; Louapre et al. 2015). Only a single 7T MRI study investigated development of cortical lesions over time (Treaba et al. 2019). That study showed that 81% of patients developed new cortical lesions after 1.5 years, with a yearly rate of 1.1 new cortical lesions in relapsing-remitting MS and 3.6 cortical lesions in SPMS patients (Treaba et al. 2019).

Twenty-five studies reported supratentorial or whole brain cortical lesion counts. Cortical lesions were frequently evident on 7T MRI, with the majority of studies reporting cortical lesions in at least 90% of patients (Mehndiratta et al. 2020; Ighani et al. 2020; Harrison, Roy, et al. 2015b; Granberg et al. 2017). Mean lesion counts per patient varied substantially across 7T MRI studies, ranging from a mean of 2.5 to 78.5 cortical lesions per patient (figure 4A). The average cortical lesion number/patient across all studies was 17 (95% CI [11-24]). Variability in 7T MRI lesion counts can at least in part be attributed to differences in the imaging sequences (see section 3.5) and differences in the clinical characteristics of the studied populations.

**Figure 4.**
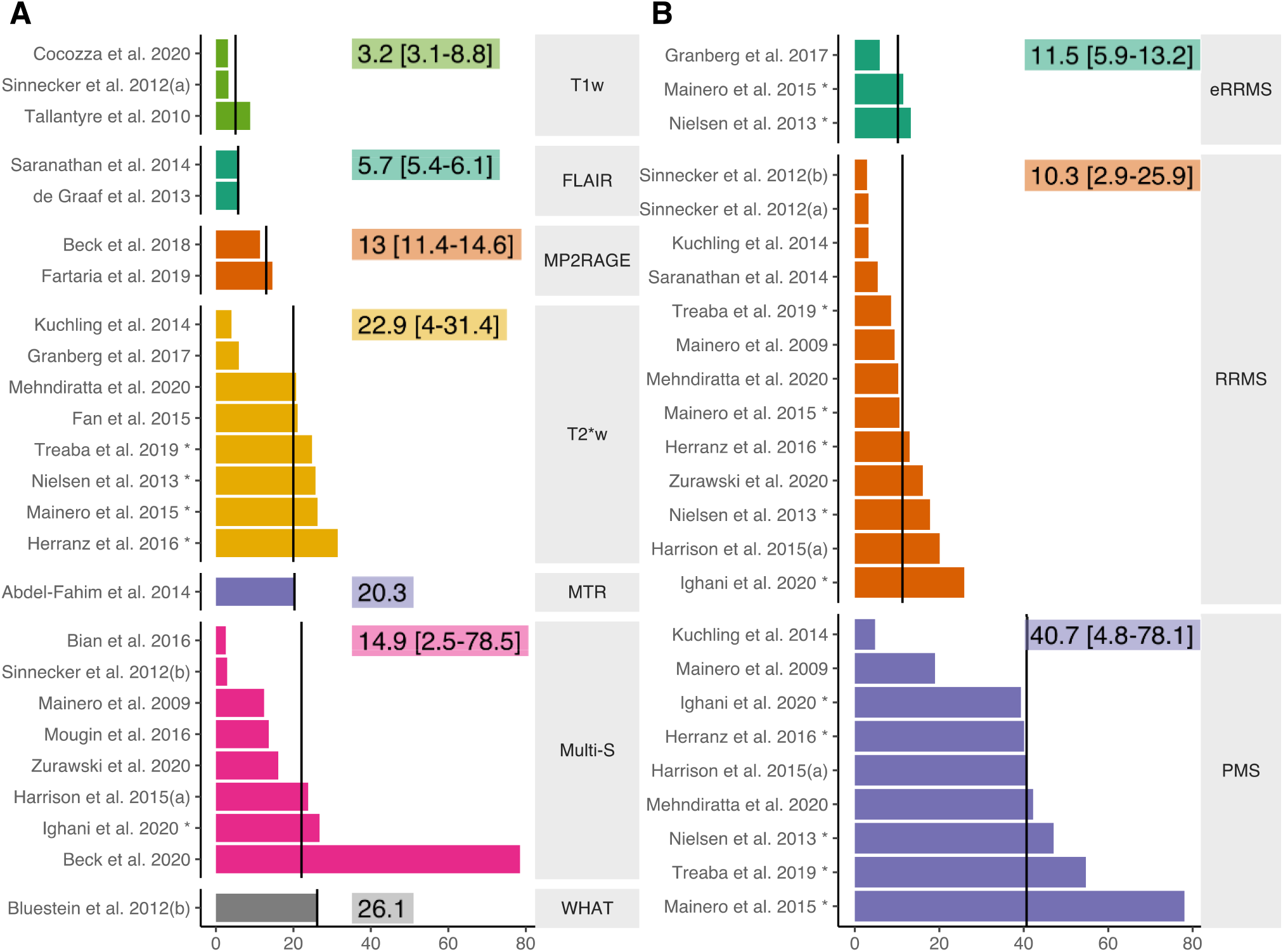
Mean cortical lesion counts per patient. **A)** 7T cortical lesion counts of the best performing sequence reported by each study, sorted by sequence. **B)** 7T cortical lesion counts per phenotype. Black vertical lines denote the mean cortical lesion count per sequence, numbers denote median and range. * = mean was estimated based on median calculations as proposed by Luo et al. (2018). Abbreviations: T1w = T1-weighted, T2*w = T2*-weighted, MTR = magnetization transfer ratio, FLAIR = fluid attenuated inversion recovery, Multi-S = multiple sequences, MP2RAGE = magnetization prepared 2-rapid gradient echo, WHAT = white matter signal attenuation, RRMS = relapsing remitting multiple sclerosis, eRRMS = early RRMS, PMS = progressive multiple sclerosis.

14 studies reported counts of cortical lesions from one or more phenotypes. PMS patients showed consistently higher cortical lesion load than RRMS patients (Cocozza et al. 2020; Harrison, Roy, et al. 2015b; Mainero et al. 2009; Mainero et al. 2015; Maranzano, Dadar, et al. 2019). Across all applied sequences, the mean lesion count was approximately four times higher in PMS (PPMS/SPMS) (41, 95% CI [27–55], n=9) compared to eRRMS (10, 95% CI [6-15], n=3) and RRMS (11, 95% CI [7–15], n=13) (figure 4B). This difference is unlikely explained by disease duration alone since cortical lesion load was found not to correlate with disease duration in individual studies (Abdel-Fahim et al. 2014; Datta et al. 2017; Mainero et al. 2009; Maranzano, Dadar, et al. 2019). Taken together, 7T MRI data seem to be in line with *post-mortem* observations, i.e. showing a marked increase in cortical lesion loads and counts in cohorts that have entered the progressive stages of MS.

### 3.4 Cortical lesion type classification at 7T

*In vivo* cortical lesion-type distributions, by 7T MRI sequence and disease phenotype, are displayed in figure 5 and 6. Corresponding lesion-type distributions from *post- mortem* studies are shown in figure 6B. 24 studies, included in the *in vivo* mean cortical lesion count analysis, reported counts of cortical lesion subtypes at 7T. On average, 63% of detected cortical lesions were classified as type I (95% CI [57%-70%]) and 37% as type II-IV lesions (95% CI [31- 43%]) (figure 5A). 12 studies reported separate counts for type II and type III/IV lesions. In these studies, 54% (95% CI [46-62%]) of lesions were type I, 11% (95% CI [7-14%]) type II and 35% (95% CI [25-46%]) type III/IV (figure 5B & 6C-E). Interestingly, we did not find any differences in lesion type ratios between RRMS and PMS groups (46% type II-IV both), while the eRRMS group had 69% type II- IV lesions (figure 5C). However, only results from three studies were included in the eRRMS group.

**Figure 5.**
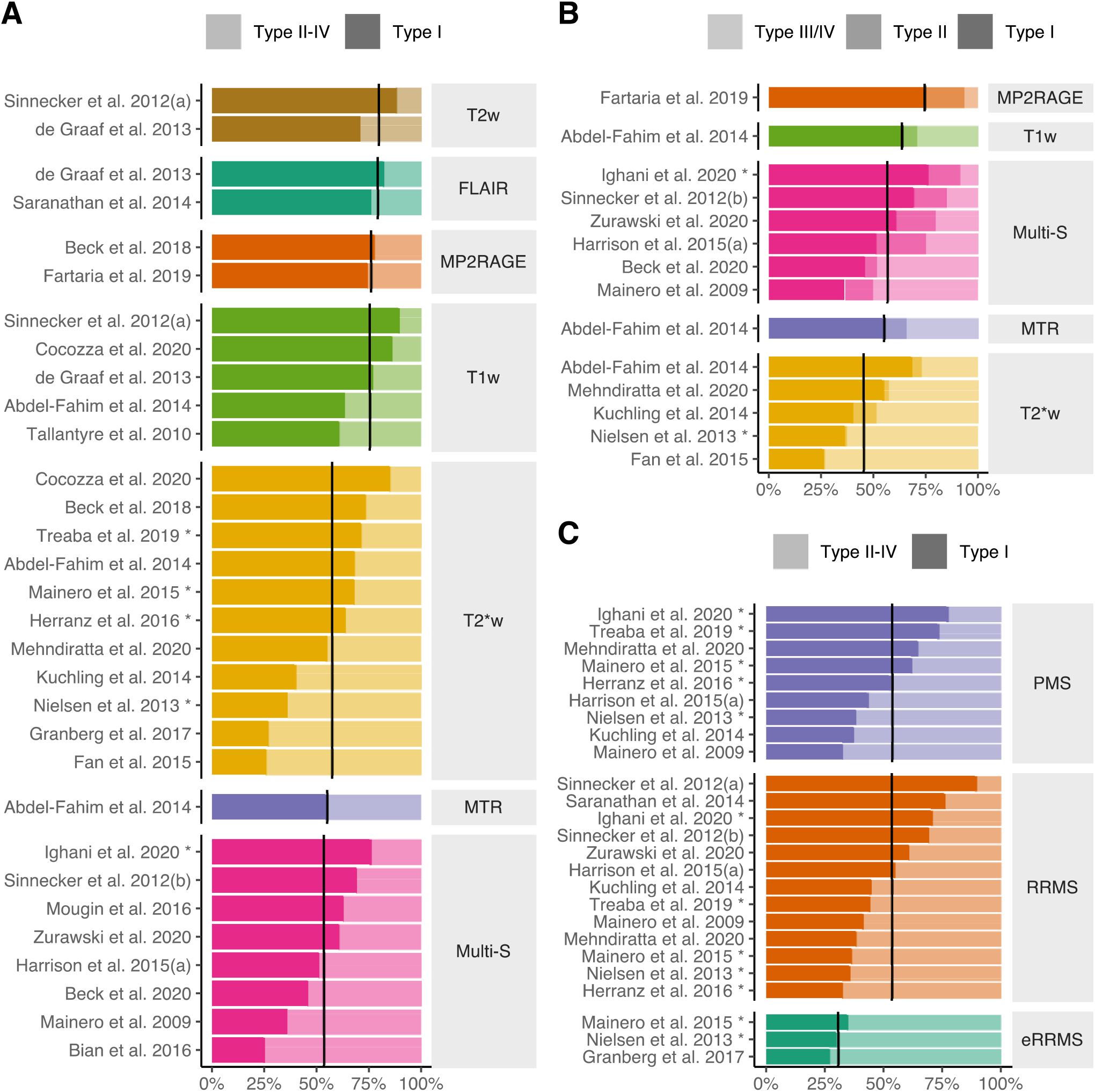
Lesion type distributions detected with 7T MRI. **A)** Lesion type ratio between type I, leukocortical, and type II-IV, intracortical lesions, for each study sorted by sequence. High opacity denotes type I lesions. **B)** Lesion type ratio between type I, type II and type III/IV for each study reporting separate type II and type III/IV lesion counts. High opacity denotes type I lesions, medium opacity denotes type II and low opacity type III/IV. **C)** Lesion type ratios for the three MS phenotypes. Denotation is the same as in A. Abbreviations: T1w = T1-weighted, T2*w = T2*-weighted, MTR = magnetization transfer ratio, FLAIR = fluid attenuated inversion recovery, T2w = T2-weighted, Multi-S: Multiple sequences, MP2RAGE: Magnetization prepared 2-rapid gradient echo, PMS: progressive multiple sclerosis, RRMS: relapsing remitting multiple sclerosis, eRRMS: early RRMS.

**Figure 6.**
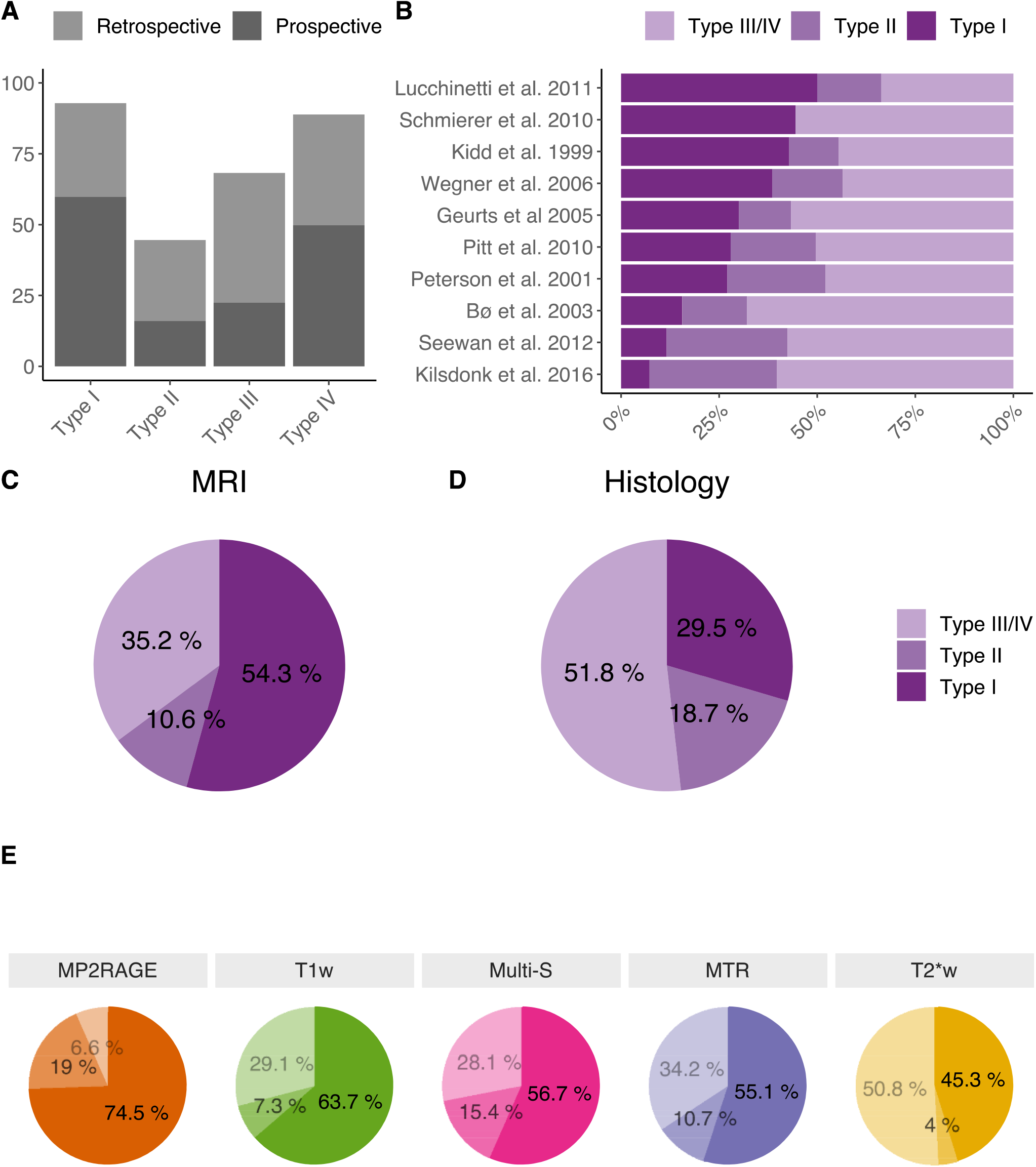
Lesion type distributions from 7T MRI and selected histological studies. **A)** Mean prospective and retrospective sensitivity of cortical lesion detection with ultra-high field MRI compared to histological staining. **B)** Cortical lesion type distributions from selected *post-mortem* studies. **C)** Mean cortical lesion type distribution from included 7T MRI studies and **D)** selected *post-mortem* studies. **E)** Mean lesion type distribution for different 7T sequences. Abbreviations: T1w = T1-weighted, T2*w = T2*-weighted, MTR = magnetization transfer ratio, FLAIR = fluid attenuated inversion recovery, T2w = T2-weighted, Multi-S: Multiple sequences, MP2RAGE: Magnetization prepared 2-rapid gradient echo, DIR = double inversion recovery, WHAT = white matter signal attenuation.

Lesion type distributions differed depending on the 7T images used for lesion segmentation and classification. FLAIR, T2w and MPRAGE/MP2RAGE identified ∼75% of lesions as leukocortical (de Graaf et al. 2013; Kilsdonk et al. 2013; Tallantyre et al. 2010; Cocozza et al. 2020). T2*w images have been suggested to be more sensitive to intracortical and subpial lesions, reporting lesion type distributions similar to histopathology (Mainero et al. 2009). From studies using either a T2*w sequence alone for lesion segmentation and classification or in combination with other MRI sequences (n=16), we found that 54% (95% CI [44-64%]) were type I and 46% (95% CI [36-55%]) were type II-IV lesions. The variation across studies might be explained by the different MS phenotypes included in these studies. Restricting the analysis to PMS patients (n=8), however, did not change the lesion type distribution.

### 3.5 Cortical lesion detection by different MRI sequences at 7T

In one of the first studies comparing multiple sequences at 7T, de Graaf et al. (2012) found no difference in cortical lesion detection between 7T FLAIR and 7T DIR, although the DIR sequence has been shown to be superior at 3T (Bouman et al. 2020; Nelson et al. 2007). However, in an expanded cohort, FLAIR detected 89% more (primarily type I) cortical lesions than DIR at 7T (Kilsdonk et al. 2013). In both studies, FLAIR and DIR detected more cortical lesions than either 2D T2w or 3D T1w, possibly due to better contrast to noise ratios between lesions and normal appearing gray matter (NAGM). Another popular sequence for cortical lesion detection is a 2D fast low-angle shot (FLASH) T2*w sequence with a high in-plane resolution (typically 0.33×0.33×1mm^3^)(Mainero et al. 2009). The T2*w magnitude image provides good gray matter/white matter contrast and a better lesion/NAGM contrast than T2w, T1w and the phase image from the FLASH sequence (Mainero et al. 2009). Because T2*w also provides excellent interrater agreement, it has been suggested as the new ‘gold standard’ for cortical lesion detection at 7T (Nielsen et al. 2012). However, 7T *post-mortem* verification studies have not confirmed superiority of T2*w, but have conversely shown either superior or similar prospective detection rates of T2w and FLAIR compared with T2*w (Jonkman et al. 2015). In addition, drawbacks of this T2*w sequence are the need to acquire 2- 3 slabs, a long total scan time of almost 20 minutes to cover the supratentorial brain (Mainero et al. 2009), and the 2D nature of the acquisition, which may hamper accurate co-registration to other image contrasts (Moraal et al. 2008). By decreasing the spatial resolution to 0.7 mm in-plane, scan time can be reduced to approximately 9 minutes, although this might influence sensitivity (Cocozza et al. 2020).

We found no *in vivo* studies that directly compared lesion detection performance of FLAIR with T2*w sequences at 7T. Only one study compared MP2RAGE with T2*w, and found T2*w performance to be inferior, revealing only half as many cortical lesions as MP2RAGE (Beck et al. 2018). Another study compared a 3D T2*w sequence at 0.5 mm isotropic resolution with an MPRAGE sequence and magnetization transfer ratio (MTR) imaging at 7T. Again the T2*w sequence performed the worst out of the three (Abdel-Fahim et al. 2014).

From the eight studies performing head-to-head sequence comparisons, FLAIR, MTR and MP2RAGE seem to outperform other sequences at 7T (table 1 & 2). Since the number of studies comparing similar sequences is small, no definitive conclusions can be drawn from the existing data. The highest mean number of cortical lesions was found in studies using either T2*w, White matter attenuation (WHAT), MTR or multiple sequences (Multi-S)(figure 4A). These results are, however, also highly confounded by the low number of studies, differences in study populations and lesion identification methods.

**Table 1.** Overview of studies directly comparing cortical lesion counts for different 7T sequences. Values for lesion types and absolute lesion count are means of the absolute supratentorial lesion count. The relative lesion count is calculated as the proportion of cortical lesions detected by each sequence compared to the best performing sequence within each sutdy. Abbreviations: T1w = T1- weighted, T2*w = T2*-weighted, MTR = magnetization transfer ratio, FLAIR = fluid attenuated inversion recovery, T2w = T2- weighted, Multi-S: Multiple sequences, MP2RAGE: Magnetization prepared 2-rapid gradient echo, DIR = double inversion recovery, WHAT = white matter signal attenuation.

| Author | Sequence | Type I | Type II | Type III/IV | Total lesion count | Relative lesion count |
| --- | --- | --- | --- | --- | --- | --- |
|  |  | Mean | Mean | Mean | Mean | Mean |
| Abdel-Fahim et al. 2014 | T2*w | 8.8 | 0.6 | 3.4 | 12.8 | 63.3 |
|  | T1w | 10.2 | 1.2 | 4.7 | 16.1 | 79.2 |
|  | MTR | 11.2 | 2.2 | 6.9 | 20.3 | 100.0 |
| Beck et al. 2018 | T2*w | 3.7 | 1.3 | - | 5.0 | 43.6 |
|  | MP2RAGE | 8.8 | 2.5 | - | 11.4 | 100.0 |
| Cocozza et al. 2020 | T2*w | 2.6 | 0.4 | - | 3.0 | 96.8 |
|  | T1w | 2.7 | 0.4 | - | 3.1 | 100.0 |
| Jonkman et al. 2015 | T2*w | 2.0 | 5.0 | 9.0 | 16.0 | 59.3 |
|  | T2w | 8.0 | 3.0 | 16.0 | 27.0 | 100.0 |
| Kilsdonk et al. 2013 | T1w | 1.4 | 0.4 | - | 1.8 | 30.5 |
|  | DIR | 1.9 | 1.2 | - | 3.1 | 52.5 |
|  | FLAIR | 4.8 | 1.1 | - | 5.9 | 100.0 |
|  | T2w | 2.2 | 0.9 | - | 3.1 | 52.5 |
| Kilsdonk et al. 2016 | T2*w | 3.0 | 0.0 | 21.0 | 24.0 | 82.8 |
|  | T1w | 4.0 | 2.0 | 18.0 | 24.0 | 82.8 |
|  | DIR | 5.0 | 1.0 | 16.0 | 22.0 | 75.9 |
|  | FLAIR | 4.0 | 3.0 | 19.0 | 26.0 | 89.7 |
|  | T2w | 6.0 | 3.0 | 20.0 | 29.0 | 100.0 |
| Pitt et al. 2010 | T2*w | 23.0 | 8.3 | 18.0 | 49.3 | 100.0 |
|  | WHAT | 21.3 | 4.3 | 10.3 | 36.0 | 73.0 |

**Table 2.** Summary statistics of the relative and absolute cortical lesion counts across studies that directly compared cortical lesion detection between different 7T sequences. The relative lesion count was calculated as the proportion of cortical lesions detected relative to the best performing sequence. Abbreviations: T1w = T1-weighted, T2*w = T2*-weighted, MTR = magnetization transfer ratio, FLAIR = fluid attenuated inversion recovery, T2w = T2-weighted, Multi-S: Multiple sequences, MP2RAGE: Magnetization prepared 2-rapid gradient echo, DIR = double inversion recovery, WHAT = white matter signal attenuation.

| Sequence | Relative lesion count | Total lesion count | Type I | Type II-IV |  |
| --- | --- | --- | --- | --- | --- |
|  | Mean | Mean | Mean | Mean | N |
| **DIR** | 64.2 | 12.6 | 3.5 | 9.1 | 2 |
| **T2\*w** | 69.9 | 16.4 | 6.7 | 9.8 | 7 |
| **WHAT** | 73.0 | 36.0 | 21.3 | 14.7 | 1 |
| **T2w** | 74.3 | 15.1 | 4.4 | 10.8 | 4 |
| **T1w** | 78.5 | 9.6 | 4.2 | 5.4 | 5 |
| **FLAIR** | 94.8 | 15.9 | 4.4 | 11.6 | 2 |
| **MTR** | 100.0 | 20.3 | 11.2 | 9.1 | 1 |
| **MP2RAGE** | 100.0 | 11.4 | 8.8 | 2.5 | 1 |

### 3.6 Post-mortem validation of cortical lesion detection at 7T and 9.4T

Four *post-mortem* studies investigated the prospective sensitivity of 7T MRI and five studies reported retrospective sensitivity at 7T (n=4) or 9.4T (n=1) MRI, verified by immunohistological myelin staining. An early study reported a prospective sensitivity of 46% using a 3D T2*w sequence at a resolution of 0.15×0.15×0.3mm^3^ (Pitt et al. 2010). Later, Kilsdonk et al. (2016) reported a prospective sensitivity of only 29% with T2*w, at 0.18mm isotropic resolution. Another study from the same group found a sensitivity of just 16% for the 3D T2*w and 28% for a 2D T2w sequence (Jonkman et al. 2015). In all three studies, sensitivity was highest for type I and type IV cortical lesions. Notably, scan times in these studies often exceeded two hours, far beyond clinical feasibility. In another study, 3D T2*w prospective sensitivity was found to be 46% at more clinically feasible scan-times (∼20 minutes for whole cerebral coverage) and 0.21 mm isotropic resolution (Yao et al. 2014). Mean prospective sensitivity calculated from four 7T studies across ten sequences was 33% (95% CI [27-39%]). Following retrospective assessment, including knowledge of lesion location from histology, sensitivity increased to 71% (95% CI [61-81%], five studies, 11 sequences). At very high resolution retrospective sensitivity may be even as high as 80 - 90% (Jonkman et al. 2015, Pitt et al. 2010). The highest retrospective sensitivity of 96 % was reported using 9.4T (Schmierer, Parkes, et al. 2010). Both prospective and retrospective sensitivity differed between lesion types. Sensitivity was highest for type I lesions (60% and 93%. respectively) followed by type IV (50% and 89%, respectively) and type III lesions (22% and 68%, respectively). Type II lesions were most difficult to identify on MRI (16% and 45%, respectively) (Figure 6A).

### 3.7 Clinical implications of cortical lesions

Eight studies reported relationships between cortical lesions detected at 7T and cognitive or motor dysfunction, using specific tests, or general disease-related impairments using the EDSS disability score.

#### 3.7.1 Relationship between cortical lesion load, cognitive and motor impairment

Five studies investigated 7T cortical lesions in relation to cognitive performance. Of these, four studies demonstrated negative correlations between cortical lesion count or volume and one or more metrics of cognition (Louapre et al. 2018; Cocozza et al. 2020; Harrison, Roy, et al. 2015b; Nielsen et al. 2013). Using a large battery of cognitive tests in visuospatial, learning/memory, processing speed and semantic domains, Nielsen et al. (2013) found that the number of type I lesions was most frequently associated with poor cognitive performance, followed by subpial lesions. This observation was replicated in another study of 36 patients, in which a natural log increase in total cortical lesion volume was associated with an odds ratio of 3.4 for being cognitively impaired. The odds ratio was 9.7 when analyzing type I lesion volume alone (Harrison, Roy, et al. 2015b). Leukocortical lesions together with age have also been shown to be the best predictor of cognitive performance in both the symbol digit modalities test (SDMT) and the revised brief visuospatial memory test (BVMT) (Cocozza et al. 2020). In summary, cortical MS lesions, and leukocortical lesions in particular, seem to contribute to cognitive decline in MS, although another recent study found no association between cortical lesion volume and SDMT performance (Mehndiratta et al. 2020).

Only two studies have investigated cortical lesions detected at 7T in relation to fine motor impairment (Harrison, Roy, et al. 2015b; Cocozza et al. 2020). In both studies, cortical lesion load correlated negatively with performance on the 9-hole peg test. This association was strongest for type I lesions.

#### 3.7.2 Relationships between cortical lesion load and EDSS

Eight studies investigated the relationship between cortical lesion load and the EDSS (Kurtzke 1983). Of these, six studies reported that cortical lesion load was positively correlated with EDSS (Louapre et al. 2018; Harrison, Roy, et al. 2015b; Mehndiratta et al. 2020; Nielsen et al. 2013; Mainero et al. 2009; Treaba et al. 2019). Cortical lesion subtype data were available from five of these six studies: In three studies, the association was primarily driven by subpial type III/IV lesions (Mainero et al. 2009; Mehndiratta et al. 2020; Nielsen et al. 2013), while leukocortical type I lesions were driving the relationship between cortical lesion load and the individual EDSS score in the remaining two studies (Harrison, Roy, et al. 2015b; Treaba et al. 2019). One study that did not find any relationship between cortical lesions and EDSS was limited by a small sample size (n=18) (Abdel-Fahim et al. 2014). In the other one, the relationship lost significance after correction for age and disease duration (Cocozza et al. 2020).

## 4. Discussion

Our systematic review on 7T MRI of cortical lesions in MS confirms the notion that 7T MRI is superior in detecting cortical lesions. *In vivo* MRI at 7T captures on average 52% more cortical lesions than the best performing MRI sequence at 3T. The relative improvement in lesion detection was most prominent for type II-IV lesions. *In vivo* 7T MRI reveals cortical lesions in the vast majority of patients and across clinical phenotypes, but the number of cortical lesions was very dependent on disease course. Importantly, we found a four-fold increase in cortical lesions in progressive compared to relapsing MS. Although *post-mortem* 7T MRI performs modestly compared to histopathology, *in vivo* 7T MRI may still prove sufficiently representative of cortical MS pathology.

### 4.1 3T MRI vs 7T MRI of cortical lesions in MS

Increases in the magnetic field strength provide a supra- linear increase in the signal to noise ratio for MRI (Pohmann, Speck, and Scheffler 2016). This offers higher spatial resolution and reduces partial volume effects, which is pivotal for the detection of cortical MS lesions (Pitt et al. 2010; Seewann et al. 2012). Our systematic review indicates that the detection of subpial and intracortical lesions improved more from 7T vs 3T MRI than detection of leukocortical type I lesions. Some studies have reported that cortical lesion detection more than doubles when comparing similar sequences at 7T and 3T (Kilsdonk et al. 2016; de Graaf et al. 2013). However, the gain from 7T was not as dramatic when we compared with the best 3T sequence in each study (52% overall and 73% considering the best 7T sequence). It should be noted that performance at different field strengths is difficult to compare as sequence parameters vary and scanning time at 7T is typically longer than at 3T (Kilsdonk et al. 2016, de Graaf et al. 2013, Abdel-Fahim et al. 2014, Maranzano et al. 2019). Decreasing scanning time will be of essence before 7T MRI technology can mature into clinical routine.

### 4.2 Extent and distribution of cortical lesions at 7T

Our review shows that cortical lesions detected with 7T MRI are prevalent and abundant in all MS stages. On average, we found 17 cortical lesions per patient.

Multiple studies found a skewed distribution in cortical lesion numbers, with some patients displaying very high cortical lesion loads, including one study that reported patients with more than 200 lesions (Beck et al. 2020). It has been suggested that patients with extensive cortical lesion load may belong to a different “myelo-cortical” phenotype of MS, where patients present with extensive cortical and spinal demyelination but few or no cerebral white matter lesions (Trapp et al. 2018). Our summarized data indicate that the variability in cortical involvement may be partly explained by differences in MS phenotypes among studies: Patients with PPMS or SPMS had considerably more cortical lesions than those with longstanding or early RRMS (Datta et al. 2017; Maranzano, Till, et al. 2019; Nielsen et al. 2013; Mainero et al. 2015; Maranzano et al. 2017). This is an important finding since cortical lesion burden at disease onset is associated with a higher risk of conversion to SPMS (Pisani et al. 2021; Scalfari et al. 2018). Longitudinal studies of clinically feasible 7T sequences are needed to help identify patients at risk for conversion from RRMS to SPMS.

With respect to the spatial distribution of cortical lesions, a slight preponderance of cortical lesion presence in prefrontal, parietal, temporal and cingulate areas was noted at 7T. This corroborates findings from earlier work at 3T, showing an accentuated cortical involvement of motor and cingulate areas (Calabrese et al. 2010). In good agreement with histopathology, 7T MRI showed that lesions are more commonly found in the sulci and deeply situated gyri as opposed to gyri located superficially in the cerebral convexities (Louapre et al. 2015; Mainero et al. 2009; Treaba et al. 2019; Kutzelnigg et al. 2005; Haider et al. 2016). This notion is further supported by a reported increase in T2* relaxation time as a measure of diffuse demyelination that was primarily located in the sulci (Mainero et al. 2015). Deeply located cortical regions may be more prone to demyelination, because they are putatively exposed to low CSF flow. This explanation would be consistent with the notion that cortical demyelination originates from perivascular meningeal infiltrates (Haider et al. 2016). It should be noted that 7T MRI has greater B_0_ and B_1_ field inhomogeneities than 3T MRI. This results in regional differences in terms of sensitivity to cortical lesions and may introduce systematic spatial biases in cortical lesion detection.

### 4.3 Cortical lesion morphology

Low spatial resolution has rendered it difficult to distinguish between the different lesion subtypes proposed by Bo et al. (2003a) when using 3T MRI (Maranzano, Dadar, et al. 2019). The improved grey/white matter contrast and higher spatial resolution at 7T allows to divide cortical lesions more easily into the different subtypes. Despite improved detection of intracortical and subpial lesions compared to 3T, and the relatively high sensitivity towards type IV lesions, the distribution of cortical lesion types revealed by 7T MRI still differed from that seen in histopathological studies (figure 6B-D). This discrepancy between radiological and histological findings can possibly be attributed to a misclassification of juxtacortical lesions as leukocortical lesions but also to insufficient sensitivity to subpial lesions on 7T MRI (Kilsdonk et al. 2016). Low subpial sensitivity may be due to lower myelin densities in the superficial cortical layers (Braitenberg 1962) resulting in lower lesion/cortex contrast, but partial volume effects and artifacts are also still present at 7T. Estimating whether *in vivo* MRI may reflect subpial type III/IV pathology is of clinical importance since subpial demyelination is an MS-specific pathology and a potential hallmark of a progressive disease course. The limited sensitivity described by *post-mortem* 7T MRI does not preclude further exploration of subpial demyelination and cortical lesions *in vivo*, since i) tissue fixation alters MRI contrasts (Schmierer, Thavarajah, et al. 2010) ii) *post-mortem* cohorts are typically biased toward longstanding and severe disease and iii) tissue sample bias may skew histological (and radiological) assessments toward more severely affected tissue. For instance, biopsy samples taken from early RRMS patients suggest that type I lesions are most preponderant earlier in the disease (Lucchinetti et al. 2011). Although this has recently been disputed (Bevan et al. 2018), longitudinal 7T MRI studies of older populations seem warranted to gain knowledge of cortical lesion morphology over time. Nevertheless, our data supports the notion that 7T MRI has the potential to visualize cortical changes associated with conversion from RRMS to SPMS and to some extent represent all cortical lesion types. Thus, subpial cortical lesion detection is no longer exclusive to histopathology, but can be reliably visualized using 7T MRI, although sensitivity towards type III lesions is still low.

### 4.4 Which MRI sequences at 7T are most suited for cortical lesion detection?

Recommending an optimal combination of 7T MRI sequences for detection of cortical lesions in MS remains difficult. Formal head-to-head comparisons remain sparse and we found high variability in cortical lesion detection between studies, even for comparable MRI sequences. Despite the low sample size and potential population sample biases, the existing data on head-to-head comparisons point toward 7T FLAIR as a reliable standard sequence for cortical lesion detection. FLAIR is widely available and can be acquired in just over five minutes at 7T (Saranathan et al. 2014). However, FLAIR suffers from B_1_ inhomogeneities at 7T, providing potentially limited applicability for whole brain assessments. In one study MTR and in another MP2RAGE appeared superior to T1w, T2w, and T2*w (Abdel-Fahim et al. 2014; Beck et al. 2018). MTR is thought to reflect presence and loss of both myelin and axons (Schmierer et al. 2007) and may therefore be better suited to distinguish MS lesions from other etiologies. MP2RAGE is a relatively new technique that provides improved gray/white matter contrast by eliminating B_0_ and B_1_ inhomogeneities, in addition to eliminating T2* signal decay. However, availability of both of these sequences for 7T MRI is still limited.

Although 7T FLAIR may be superior for detection of cortical lesions in general, and hence for monitoring clinically definite MS patients, 7T 2D-T2*w may be the best current sequence for detection of subpial type III/VI lesions, and hence for diagnosis of MS, including PPMS or MOG-AD. Timely identification of subpial lesions by T2*w could prove to accelerate the stratification of RRMS patients at risk for conversion to SPMS. While some leukocortical lesions may go undetected by 2D-T2*w (Maranzano, Dadar, et al. 2019), this might be compensated by including FLAIR. Therefore, the combination of FLAIR and 2D-T2*w sequences might offer a feasible protocol that reliably reflects developments in number and volume of cortical lesions and their individual subtypes.

Parallel to improving existing 7T sequences, novel sequences are being developed to improve contrast for cortical lesions. WHAT T1w (Bluestein, Pitt, Sammet, et al. 2012b; Saranathan et al. 2015), is a promising tool for improving detection of especially type I lesions (Bluestein, Pitt, Sammet, et al. 2012b). More recently, PSIR, popular for visualization of cortical lesions at 3T (Bouman et al. 2020), has been adapted to 7T by including a *null point image* (NPI) into the MP2RAGE sequence (Mougin et al. 2016), yielding another promising avenue for future improvements. However, more head-to-head comparisons with existing sequences at 7T are needed in order to assess the advantages of these new sequences.

### 4.5 Segmentation and classification of cortical lesions

Standardized identification and segmentation of cortical lesions is of pivotal importance in studies assessing the value of cortical lesions in diagnosis and monitoring of MS. Whereas guidelines for segmentation and scoring of cortical lesions have been established for 1.5T and 3T DIR (Geurts et al. 2011), few recommendations exist for 7T. Our synthesis showed that DIR guidelines sometimes are applied to 7T MRI data (de Graaf et al. 2013; Kilsdonk et al. 2013; Saranathan et al. 2014). However, given the higher resolution of anatomical scans at 7T, adhering to these guidelines may lead to segmentation of smaller lesions and potentially increase the false-positive detection rate. Different sequence- or field-strength related artifacts, including B_0_ and B_1_ inhomogeneities, could complicate application across field strengths even further. Moreover, we found that the procedure and criteria for cortical lesion segmentation were only reported in 32 and 28 studies, respectively. This complicates study replication and impairs the training of less experienced readers. It is very likely that the large variability observed in cortical lesion counts between studies (see figure 4) is in part due to differences in segmentation procedures. Thus, there is a pressing need for the development of guidelines directed at segmentation and scoring of cortical lesions at 7T.

### 4.6 Clinical impact of cortical lesions

At 1.5T and 3T, cortical lesions have been related to both clinical severity and disease progression (Calabrese et al. 2012; Scalfari et al. 2018; Nelson et al. 2011; Mike et al. 2011; Forslin et al. 2018). However, as cortical lesion detection depends on lesion size and myelin density in the surrounding tissue, MRI at 3T and 1.5T preferentially detects leukocortical type I lesions (Forslin et al. 2018; Mike et al. 2011). Consequently, results from 3T and 1.5T MRI studies might underestimate how clinical severity is influenced by cortical lesions, and subpial lesions in particular.

Our study review supports the notion that cortical lesions are a highly relevant marker of clinical severity, relating to both cognitive and motor function. Associations between specific cortical lesion types and disability measures were less clear, and might be biased by the MRI sequence used, e.g. differential strength of T2*w vs FLAIR at 7T for detecting different aspects of cortical lesion formation. Episodes of cognitive worsening have been documented in relation to cortical lesion development at 3T, supporting a notion of “cortical” relapses due to newly developing cortical lesions (Puthenparampil et al. 2016). Rather than focusing on whole brain lesion counts and overall clinical impairment, future studies should address the structure-function relations between the manifestation of individual cortical lesions, in terms of location, lesion-type, and impairment within specific functional domains, such as motor, sensory, cognitive impairment or fatigue. Mental fatigue is one of the most frequent and disabling symptoms in MS. Clinicians and patients are frequently frustrated by this symptom, which cannot be accounted for by white matter lesions alone (Sepulcre et al. 2009). An impact of cortical lesions on fatigue is plausible, since neurons in cortical MS lesions show signs of mitochondrial injury and respiratory chain dysfunction (Campbell, Worrall, and Mahad 2014). However, assessment of the independent contribution of cortical lesions to impairment of specific domains is complicated by confounding factors, including e.g. atrophy and spinal cord involvement. Future study designs should account or correct for these factors by patient selection in longitudinal studies or by advanced statistical procedures in cross-sectional designs.

### 4.7 Ultra-high field MRI in the clinical setting

Despite the well-documented advantages of 7T MRI in MS, it has not yet been implemented as a clinical tool. Fortunately, the recent certifications of some 7T MRI systems by the Food and Drug Administration (FDA) and the Conformité Européenne (CE), may facilitate clinical use of 7T MRI in MS. Using the MAGNIMS recommended protocol for clinical workup (T1w, T2w and FLAIR images) (Rovira et al. 2015), 7T images acquired in ∼30 minutes improved cortical lesion detection by all three sequences compared to corresponding 3T sequences without compromising sensitivity for white matter lesions (de Graaf et al. 2013). Furthermore, in a clinically feasible protocol, including T2*w at a total scan time of ∼22 minutes, all cortical lesion types were captured with high inter-rater agreement in patients with RRMS and PMS (Cocozza et al. 2020). One study comparing 7T with 3T MRI in a clinical setting described increased confidence in MS diagnosis with 7T MRI, which related partly to improved cortical lesion detection (Springer et al. 2016). Additionally, histopathological data indicated that 7T may accelerate differential diagnosis of primary demyelinating diseases such as MS and rare acute syndromes associated with IgG antibodies against myelin oligodendrocyte glycoprotein as opposed to other diseases with features of demyelination such as neuromyelitis optica spectrum disorder, hereditary leukodystrophies and neuro-infections (Sinnecker, Dorr, et al. 2012b; Behrens et al. 2018; Junker et al. 2020; Fischer et al. 2013). However, the potential for improved differential diagnosis needs confirmation by systematic comparative studies at 7T. A thorough review on the diagnostic potential of 7T MRI in MS can be found elsewhere (Sati 2018). There are, however, also technical challenges with 7T MRI, including higher B_1_ and B_0_ field inhomogeneities, potentially greater impact of subject motion and specific absorption rate limitations that need to be addressed before 7T can become a useful clinical tool (Sati 2018). In addition, stricter safety precautions may exclude more patients from 7T as compared to 3T MRI.

### 4.8 Limitations

The conversion from median and range of cortical lesion counts to means from some studies limits the interpretation of our results. Cortical lesion counts often have a skewed distribution, biasing the estimations of group means, although the method used in this study provides a better protection against this bias than previous ones (Luo et al. 2018). We did not perform any statistical testing due to the low number of studies included in the sub-analyses. Therefore, the numbers are not corrected for variations in population demographics, which might limit the interpretability of our results. Because many studies only reported combined intracortical lesion counts, we also pooled cortical lesion type II and III/IV into one category, although histopathology proposes that the two lesion types have different origins (Peterson et al. 2001; Kidd et al. 1999). Pooling may be justified in so far as type II-IV lesions are all drained by strictly intracortical venules as opposed to type I lesions, which are associated with principal cortical veins that are transcortical and drain into the juxta-cortical white matter (Duvernoy, Delon, and Vannson 1981; Kidd et al. 1999). However, compared to type II and type I lesions, type III/IV subpial lesions may be more specific for MS or MOG-AD.

## 5. Conclusion

7T MRI improves cortical lesion detection in MS compared to clinical field strengths, especially for subpial demyelination which is of high diagnostic and prognostic relevance. However, our review emphasizes the need for standardization in both acquisition and processing (i.e. segmenting) of 7T MRI data. Nonetheless, based on the combined findings from 7T MRI studies, is seems that the MAGNIMS protocol at 7T is clinically feasible and improves detection and monitoring of cortical lesions. Adding a sequence more sensitive to subpial lesions, like T2*w, would most likely be beneficial, although further histopathological validation is warranted. Importantly, improved cortical lesion visualization at 7T can be achieved without compromising the sensitivity for white matter lesions. By providing a clearer view of cortical pathology, 7T will improve the management of demyelinating diseases by accelerating differential diagnosis of encephalopathies affecting myelin and phenotyping within the MS-spectrum. Alleviating the difficulty in MS phenotyping is of immense clinical concern since emerging therapies are licensed and accessible only to patients of specific MS phenotypes.

## Data Availability

Data used in the article is freely available from previously published papers.

## Abbreviations

MS: multiple sclerosis
UHF: ultra-high field
CIS: clinically isolated syndrome
RRMS: relapsing remitting multiple sclerosis
CNS: central nervous system
MRI: magnetic resonance imaging
MOG-AD: anti-MOG-IgG
SPMS: secondary progressive multiple sclerosis
PPMS: primary progressive multiple sclerosis
EAE: experimental autoimmune encephalomyelitis
DIR: double inversion recovery
CSF: cerebrospinal fluid
FLAIR: fluid attenuated inversion recovery
PSIR: phase sensitive inversion recovery
IR-SWIET: inversion recovery susceptibility weighted imaging with enhanced T2 weighting
PRISMA: preferred reporting items for systematic reviews and meta-analyses
EDSS: expanded disability status scale
eRRMS: early relapsing remitting multiple sclerosis
PMS: progressive multiple sclerosis
w: weighted
MP2RAGE: magnetization prepared 2 rapid gradient echo
NAGM: normal appearing gray matter
FLASH: fast low-angle shot
MTR: magnetization transfer ratio
WHAT: white matter attenuation
SDMT: symbol digit modalities test
BVMT: brief visuospatial memory test
NPI: null point image

## References

Abdel-Fahim, R., N. Mistry, O. Mougin, A. Blazejewska, A. Pitiot, R. Retkute, P. Gowland, and N. Evangelou. 2014. ‘Improved detection of focal cortical lesions using 7T magnetisation transfer imaging in patients with multiple sclerosis’, Mult Scler Relat Disord, 3: 258–65.

Barkhof, F. 1999. ‘MRI in multiple sclerosis: correlation with expanded disability status scale (EDSS)’, Mult Scler, 5: 283–6.

Beck, E. S., N. Gai, S. Filippini, J. Maranzano, G. Nair, and D. S. Reich. 2020. ‘Inversion Recovery Susceptibility Weighted Imaging With Enhanced T2 Weighting at 3 T Improves Visualization of Subpial Cortical Multiple Sclerosis Lesions’, Invest Radiol, 55: 727–35.

Beck, E. S., P. Sati, V. Sethi, T. Kober, B. Dewey, P. Bhargava, G. Nair, I. C. Cortese, and D. S. Reich. 2018. ‘Improved Visualization of Cortical Lesions in Multiple Sclerosis Using 7T MP2RAGE’, AJNR Am J Neuroradiol, 39: 459–66.

Behrens, J. R., J. Wanner, J. Kuchling, L. Ostendorf, L. Harms, K. Ruprecht, T. Niendorf, S. Jarius, B. Wildemann, R. M. Giess, M. Scheel, J. Bellmann-Strobl, J. Wuerfel, F. Paul, and T. Sinnecker. 2018. ‘7 Tesla MRI of Balo’s concentric sclerosis versus multiple sclerosis lesions’, Ann Clin Transl Neurol, 5: 900–12.

Bevan, R. J., R. Evans, L. Griffiths, L. M. Watkins, M. I. Rees, R. Magliozzi, I. Allen, G. McDonnell, R. Kee, M. Naughton, D. C. Fitzgerald, R. Reynolds, J. W. Neal, and O. W. Howell. 2018. ‘Meningeal inflammation and cortical demyelination in acute multiple sclerosis’, Ann Neurol, 84: 829–42.

Bian, W., E. Tranvinh, T. Tourdias, M. Han, T. Liu, Y. Wang, B. Rutt, and M. M. Zeineh. 2016. ‘In Vivo 7T MR Quantitative Susceptibility Mapping Reveals Opposite Susceptibility Contrast between Cortical and White Matter Lesions in Multiple Sclerosis’, AJNR Am J Neuroradiol, 37: 1808–15.

Bluestein, K. T., D. Pitt, M. V. Knopp, and P. Schmalbrock. 2012a. ‘T1 and proton density at 7 T in patients with multiple sclerosis: an initial study’, Magn Reson Imaging, 30: 19–25.

Bluestein, K. T., D. Pitt, S. Sammet, C. R. Zachariah, U. Nagaraj, M. V. Knopp, and P. Schmalbrock. 2012b. ‘Detecting cortical lesions in multiple sclerosis at 7 T using white matter signal attenuation’, Magn Reson Imaging, 30: 907–15.

Bø, L., C. A. Vedeler, H. I. Nyland, B. D. Trapp, and S. J. Mork. 2003a. ‘Subpial demyelination in the cerebral cortex of multiple sclerosis patients’, J Neuropathol Exp Neurol, 62: 723–32.

Bø, L., C. A. Vedeler, H. Nyland, B. D. Trapp, and S. J. Mork. 2003b. ‘Intracortical multiple sclerosis lesions are not associated with increased lymphocyte infiltration’, Mult Scler, 9: 323–31.

Bouman, P. M., M. D. Steenwijk, P. J. W. Pouwels, M. M. Schoonheim, F. Barkhof, L. E. Jonkman, and J. J. G. Geurts. 2020. ‘Histopathology-validated recommendations for cortical lesion imaging in multiple sclerosis’, Brain.

Braitenberg, V. 1962. ‘A note on myeloarchitectonics’, J Comp Neurol, 118: 141–56.

Brownell, B., and J. T. Hughes. 1962. ‘The distribution of plaques in the cerebrum in multiple sclerosis’, J Neurol Neurosurg Psychiatry, 25: 315–20.

Calabrese, M., M. Battaglini, A. Giorgio, M. Atzori, V. Bernardi, I. Mattisi, P. Gallo, and N. De Stefano. 2010. ‘Imaging distribution and frequency of cortical lesions in patients with multiple sclerosis’, Neurology, 75: 1234–40.

Calabrese, M., V. Poretto, A. Favaretto, S. Alessio, V. Bernardi, C. Romualdi, F. Rinaldi, P. Perini, and P. Gallo. 2012. ‘Cortical lesion load associates with progression of disability in multiple sclerosis’, Brain, 135: 2952–61.

Campbell, G. R., J. T. Worrall, and D. J. Mahad. 2014. ‘The central role of mitochondria in axonal degeneration in multiple sclerosis’, Mult Scler, 20: 1806–13.

Choi, S. R., O. W. Howell, D. Carassiti, R. Magliozzi, D. Gveric, P. A. Muraro, R. Nicholas, F. Roncaroli, and R. Reynolds. 2012. ‘Meningeal inflammation plays a role in the pathology of primary progressive multiple sclerosis’, Brain, 135: 2925–37.

Cocozza, S., M. Cosottini, A. Signori, L. Fleysher, M. M. El Mendili, F. Lublin, M. Inglese, and L. Roccatagliata. 2020. ‘A clinically feasible 7-Tesla protocol for the identification of cortical lesions in Multiple Sclerosis’, Eur Radiol, 30: 4586–94.

Cohen-Adad, J., T. Benner, D. Greve, R. P. Kinkel, A. Radding, B. Fischl, B. R. Rosen, and C. Mainero. 2011. ‘In vivo evidence of disseminated subpial T2* signal changes in multiple sclerosis at 7 T: a surface-based analysis’, Neuroimage, 57: 55–62.

Datta, R., V. Sethi, S. Ly, A. T. Waldman, S. Narula, B. E. Dewey, P. Sati, D. Reich, and B. Banwell. 2017. ‘7T MRI Visualization of Cortical Lesions in Adolescents and Young Adults with Pediatric-Onset Multiple Sclerosis’, J Neuroimaging, 27: 447–52.

Dawson, J. W. 1916. ‘The histology of disseminated sclerosis’, Trans Roy Soc Edinb, 50: 517–740.

de Graaf, W. L., I. D. Kilsdonk, A. Lopez-Soriano, J. J. Zwanenburg, F. Visser, C. H. Polman, J. A. Castelijns, J. J. Geurts, P. J. Pouwels, P. R. Luijten, F. Barkhof, and M. P. Wattjes. 2013. ‘Clinical application of multi-contrast 7-T MR imaging in multiple sclerosis: increased lesion detection compared to 3 T confined to grey matter’, Eur Radiol, 23: 528–40.

de Graaf, W. L., J. J. Zwanenburg, F. Visser, M. P. Wattjes, P. J. Pouwels, J. J. Geurts, C. H. Polman, F. Barkhof, P. R. Luijten, and J. A. Castelijns. 2012. ‘Lesion detection at seven Tesla in multiple sclerosis using magnetisation prepared 3D-FLAIR and 3D-DIR’, Eur Radiol, 22: 221–31.

Dury, R. J., Y. Falah, P. A. Gowland, N. Evangelou, M. G. Bright, and S. T. Francis. 2019. ‘Ultra-high-field arterial spin labelling MRI for non-contrast assessment of cortical lesion perfusion in multiple sclerosis’, Eur Radiol, 29: 2027–33.

Duvernoy, H. M., S. Delon, and J. L. Vannson. 1981. ‘Cortical blood vessels of the human brain’, Brain Res Bull, 7: 519–79.

Fan, A. P., S. T. Govindarajan, R. P. Kinkel, N. K. Madigan, A. S. Nielsen, T. Benner, E. Tinelli, B. R. Rosen, E. Adalsteinsson, and C. Mainero. 2015. ‘Quantitative oxygen extraction fraction from 7-Tesla MRI phase: reproducibility and application in multiple sclerosis’, J Cereb Blood Flow Metab, 35: 131–9.

Fartaria, M. J., K. O’Brien, A. Sorega, G. Bonnier, A. Roche, P. Falkovskiy, G. Krueger, T. Kober, M. Bach Cuadra, and C. Granziera. 2017. ‘An Ultra-High Field Study of Cerebellar Pathology in Early Relapsing-Remitting Multiple Sclerosis Using MP2RAGE’, Invest Radiol, 52: 265–73.

Fartaria, M. J., P. Sati, A. Todea, E. W. Radue, R. Rahmanzadeh, K. O’Brien, D. S. Reich, M. Bach Cuadra, T. Kober, and C. Granziera. 2019. ‘Automated Detection and Segmentation of Multiple Sclerosis Lesions Using Ultra-High-Field MP2RAGE’, Invest Radiol, 54: 356–64.

Fischer, M. T., I. Wimmer, R. Hoftberger, S. Gerlach, L. Haider, T. Zrzavy, S. Hametner, D. Mahad, C. J. Binder, M. Krumbholz, J. Bauer, M. Bradl, and H. Lassmann. 2013. ‘Disease-specific molecular events in cortical multiple sclerosis lesions’, Brain, 136: 1799–815.

Forslin, Y., A. Bergendal, F. Hashim, J. Martola, S. Shams, M. K. Wiberg, S. Fredrikson, and T. Granberg. 2018. ‘Detection of Leukocortical Lesions in Multiple Sclerosis and Their Association with Physical and Cognitive Impairment: A Comparison of Conventional and Synthetic Phase-Sensitive Inversion Recovery MRI’, AJNR Am J Neuroradiol, 39: 1995–2000.

Gaitan, M. I., P. Sati, S. J. Inati, and D. S. Reich. 2013. ‘Initial investigation of the blood-brain barrier in MS lesions at 7 tesla’, Mult Scler, 19: 1068–73.

Geurts, J. J., L. Bo, P. J. Pouwels, J. A. Castelijns, C. H. Polman, and F. Barkhof. 2005. ‘Cortical lesions in multiple sclerosis: combined postmortem MR imaging and histopathology’, AJNR Am J Neuroradiol, 26: 572–7.

Geurts, J. J., P. J. Pouwels, B. M. Uitdehaag, C. H. Polman, F. Barkhof, and J. A. Castelijns. 2005. ‘Intracortical lesions in multiple sclerosis: improved detection with 3D double inversion-recovery MR imaging’, Radiology, 236: 254–60.

Geurts, J. J., S. D. Roosendaal, M. Calabrese, O. Ciccarelli, F. Agosta, D. T. Chard, A. Gass, E. Huerga, B. Moraal, D. Pareto, M. A. Rocca, M. P. Wattjes, T. A. Yousry, B. M. Uitdehaag, F. Barkhof, and Magnims Study Group. 2011. ‘Consensus recommendations for MS cortical lesion scoring using double inversion recovery MRI’, Neurology, 76: 418–24.

Granberg, T., Q. Fan, C. A. Treaba, R. Ouellette, E. Herranz, G. Mangeat, C. Louapre, J. Cohen-Adad, E. C. Klawiter, J. A. Sloane, and C. Mainero. 2017. ‘In vivo characterization of cortical and white matter neuroaxonal pathology in early multiple sclerosis’, Brain, 140: 2912–26.

Haider, L., T. Zrzavy, S. Hametner, R. Hoftberger, F. Bagnato, G. Grabner, S. Trattnig, S. Pfeifenbring, W. Bruck, and H. Lassmann. 2016. ‘The topograpy of demyelination and neurodegeneration in the multiple sclerosis brain’, Brain, 139: 807–15.

Harrison, D. M., J. Oh, S. Roy, E. T. Wood, A. Whetstone, M. A. Seigo, C. K. Jones, D. Pham, P. van Zijl, D. S. Reich, and P. A. Calabresi. 2015a. ‘Thalamic lesions in multiple sclerosis by 7T MRI: Clinical implications and relationship to cortical pathology’, Mult Scler, 21: 1139–50.

Harrison, D. M., S. Roy, J. Oh, I. Izbudak, D. Pham, S. Courtney, B. Caffo, C. K. Jones, P. van Zijl, and P. A. Calabresi. 2015b. ‘Association of Cortical Lesion Burden on 7-T Magnetic Resonance Imaging With Cognition and Disability in Multiple Sclerosis’, JAMA Neurol, 72: 1004–12.

Herranz, E., C. Gianni, C. Louapre, C. A. Treaba, S. T. Govindarajan, R. Ouellette, M. L. Loggia, J. A. Sloane, N. Madigan, D. Izquierdo-Garcia, N. Ward, G. Mangeat, T. Granberg, E. C. Klawiter, C. Catana, J. M. Hooker, N. Taylor, C. Ionete, R. P. Kinkel, and C. Mainero. 2016. ‘Neuroinflammatory component of gray matter pathology in multiple sclerosis’, Ann Neurol, 80: 776–90.

Herranz, E., C. Louapre, C. A. Treaba, S. T. Govindarajan, R. Ouellette, G. Mangeat, M. L. Loggia, J. Cohen-Adad, E. C. Klawiter, J. A. Sloane, and C. Mainero. 2020. ‘Profiles of cortical inflammation in multiple sclerosis by (11)C-PBR28 MR-PET and 7 Tesla imaging’, Mult Scler, 26: 1497–509.

Höftberger, R., Y. Guo, E. P. Flanagan, A. S. Lopez-Chiriboga, V. Endmayr, S. Hochmeister, D. Joldic, S. J. Pittock, J. M. Tillema, M. Gorman, H. Lassmann, and C. F. Lucchinetti. 2020. ‘The pathology of central nervous system inflammatory demyelinating disease accompanying myelin oligodendrocyte glycoprotein autoantibody’, Acta Neuropathol, 139: 875–92.

Howell, O. W., C. A. Reeves, R. Nicholas, D. Carassiti, B. Radotra, S. M. Gentleman, B. Serafini, F. Aloisi, F. Roncaroli, R. Magliozzi, and R. Reynolds. 2011. ‘Meningeal inflammation is widespread and linked to cortical pathology in multiple sclerosis’, Brain, 134: 2755–71.

Hulst, H. E., and J. J. Geurts. 2011. ‘Gray matter imaging in multiple sclerosis: what have we learned?’, BMC Neurol, 11: 153.

Ighani, M., S. Jonas, I. Izbudak, S. Choi, A. Lema-Dopico, J. Hua, E. E. O’Connor, and D. M. Harrison. 2020. ‘No association between cortical lesions and leptomeningeal enhancement on 7-Tesla MRI in multiple sclerosis’, Mult Scler, 26: 165–76.

Jonkman, L. E., L. Fleysher, M. D. Steenwijk, J. A. Koeleman, T. P. de Snoo, F. Barkhof, M. Inglese, and J. J. Geurts. 2016a. ‘Ultra-high field MTR and qR2* differentiates subpial cortical lesions from normal-appearing gray matter in multiple sclerosis’, Mult Scler, 22: 1306–14.

Jonkman, L. E., R. Klaver, L. Fleysher, M. Inglese, and J. J. Geurts. 2015. ‘Ultra-High-Field MRI Visualization of Cortical Multiple Sclerosis Lesions with T2 and T2*: A Postmortem MRI and Histopathology Study’, AJNR Am J Neuroradiol, 36: 2062–7.

Jonkman, L. E., R. Klaver, L. Fleysher, M. Inglese, and J. J. Geurts. 2016b. ‘The substrate of increased cortical FA in MS: A 7T post-mortem MRI and histopathology study’, Mult Scler, 22: 1804–11.

Junker, A., J. Wozniak, D. Voigt, U. Scheidt, J. Antel, C. Wegner, W. Bruck, and C. Stadelmann. 2020. ‘Extensive subpial cortical demyelination is specific to multiple sclerosis’, Brain Pathol, 30: 641–52.

Kangarlu, A., E. C. Bourekas, A. Ray-Chaudhury, and K. W. Rammohan. 2007. ‘Cerebral cortical lesions in multiple sclerosis detected by MR imaging at 8 Tesla’, AJNR Am J Neuroradiol, 28: 262–6.

Kidd, D., F. Barkhof, R. McConnell, P. R. Algra, I. V. Allen, and T. Revesz. 1999. ‘Cortical lesions in multiple sclerosis’, Brain, 122 (Pt 1): 17–26.

Kilsdonk, I. D., W. L. de Graaf, A. L. Soriano, J. J. Zwanenburg, F. Visser, J. P. Kuijer, J. J. Geurts, P. J. Pouwels, C. H. Polman, J. A. Castelijns, P. R. Luijten, F. Barkhof, and M. P. Wattjes. 2013. ‘Multicontrast MR imaging at 7T in multiple sclerosis: highest lesion detection in cortical gray matter with 3D-FLAIR’, AJNR Am J Neuroradiol, 34: 791–6.

Kilsdonk, I. D., L. E. Jonkman, R. Klaver, S. J. van Veluw, J. J. Zwanenburg, J. P. Kuijer, P. J. Pouwels, J. W. Twisk, M. P. Wattjes, P. R. Luijten, F. Barkhof, and J. J. Geurts. 2016. ‘Increased cortical grey matter lesion detection in multiple sclerosis with 7 T MRI: a post-mortem verification study’, Brain, 139: 1472–81.

Kolber, P., A. Droby, A. Roebroeck, R. Goebel, V. Fleischer, S. Groppa, and F. Zipp. 2017. ‘A “kissing lesion”: In-vivo 7T evidence of meningeal inflammation in early multiple sclerosis’, Mult Scler, 23: 1167–69.

Kollia, K., S. Maderwald, N. Putzki, M. Schlamann, J. M. Theysohn, O. Kraff, M. E. Ladd, M. Forsting, and I. Wanke. 2009. ‘First clinical study on ultra-high-field MR imaging in patients with multiple sclerosis: comparison of 1.5T and 7T’, AJNR Am J Neuroradiol, 30: 699–702.

Kuchling, J., C. Ramien, I. Bozin, J. Dorr, L. Harms, B. Rosche, T. Niendorf, F. Paul, T. Sinnecker, and J. Wuerfel. 2014. ‘Identical lesion morphology in primary progressive and relapsing-remitting MS--an ultrahigh field MRI study’, Mult Scler, 20: 1866–71.

Kurtzke, J. F. 1983. ‘Rating neurologic impairment in multiple sclerosis: an expanded disability status scale (EDSS)’, Neurology, 33: 1444–52.

Kutzelnigg, A., C. F. Lucchinetti, C. Stadelmann, W. Bruck, H. Rauschka, M. Bergmann, M. Schmidbauer, J. E. Parisi, and H. Lassmann. 2005. ‘Cortical demyelination and diffuse white matter injury in multiple sclerosis’, Brain, 128: 2705–12.

Lassmann, H. 2005. ‘Multiple sclerosis pathology: evolution of pathogenetic concepts’, Brain Pathol, 15: 217–22.

Louapre, C., S. T. Govindarajan, C. Gianni, C. Langkammer, J. A. Sloane, R. P. Kinkel, and C. Mainero. 2015. ‘Beyond focal cortical lesions in MS: An in vivo quantitative and spatial imaging study at 7T’, Neurology, 85: 1702–9.

Louapre, C., S. T. Govindarajan, C. Gianni, N. Madigan, J. A. Sloane, C. A. Treaba, E. Herranz, R. P. Kinkel, and C. Mainero. 2018. ‘Heterogeneous pathological processes account for thalamic degeneration in multiple sclerosis: Insights from 7 T imaging’, Mult Scler, 24: 1433–44.

Lucchinetti, C. F., B. F. Popescu, R. F. Bunyan, N. M. Moll, S. F. Roemer, H. Lassmann, W. Bruck, J. E. Parisi, B. W. Scheithauer, C. Giannini, S. D. Weigand, J. Mandrekar, and R. M. Ransohoff. 2011. ‘Inflammatory cortical demyelination in early multiple sclerosis’, N Engl J Med, 365: 2188–97.

Luo, D., X. Wan, J. Liu, and T. Tong. 2018. ‘Optimally estimating the sample mean from the sample size, median, mid-range, and/or mid-quartile range’, Stat Methods Med Res, 27: 1785–805.

Magliozzi, R., O. Howell, A. Vora, B. Serafini, R. Nicholas, M. Puopolo, R. Reynolds, and F. Aloisi. 2007. ‘Meningeal B-cell follicles in secondary progressive multiple sclerosis associate with early onset of disease and severe cortical pathology’, Brain, 130: 1089–104.

Mainero, C., T. Benner, A. Radding, A. van der Kouwe, R. Jensen, B. R. Rosen, and R. P. Kinkel. 2009. ‘In vivo imaging of cortical pathology in multiple sclerosis using ultra-high field MRI’, Neurology, 73: 941–8.

Mainero, C., C. Louapre, S. T. Govindarajan, C. Gianni, A. S. Nielsen, J. Cohen-Adad, J. Sloane, and R. P. Kinkel. 2015. ‘A gradient in cortical pathology in multiple sclerosis by in vivo quantitative 7 T imaging’, Brain, 138: 932–45.

Maranzano, J., M. Dadar, D. A. Rudko, D. De Nigris, C. Elliott, J. S. Gati, S. A. Morrow, R. S. Menon, D. L. Collins, D. L. Arnold, and S. Narayanan. 2019. ‘Comparison of Multiple Sclerosis Cortical Lesion Types Detected by Multicontrast 3T and 7T MRI’, AJNR Am J Neuroradiol, 40: 1162–69.

Maranzano, J., D. A. Rudko, D. L. Arnold, and S. Narayanan. 2016. ‘Manual Segmentation of MS Cortical Lesions Using MRI: A Comparison of 3 MRI Reading Protocols’, AJNR Am J Neuroradiol, 37: 1623–8.

Maranzano, J., D. A. Rudko, K. Nakamura, S. Cook, D. Cadavid, L. Wolansky, D. L. Arnold, and S. Narayanan. 2017. ‘MRI evidence of acute inflammation in leukocortical lesions of patients with early multiple sclerosis’, Neurology, 89: 714–21.

Maranzano, J., C. Till, H. E. Assemlal, V. Fonov, R. Brown, D. Araujo, J. O’Mahony, E. A. Yeh, A. Bar-Or, R. A. Marrie, L. Collins, B. Banwell, D. L. Arnold, and S. Narayanan. 2019. ‘Detection and clinical correlation of leukocortical lesions in pediatric-onset multiple sclerosis on multi-contrast MRI’, Mult Scler, 25: 980–86.

Mehndiratta, A., C. A. Treaba, V. Barletta, E. Herranz, R. Ouellette, J. A. Sloane, E. C. Klawiter, R. P. Kinkel, and C. Mainero. 2020. ‘Characterization of thalamic lesions and their correlates in multiple sclerosis by ultra-high-field MRI’, Mult Scler: 1352458520932804.

Merkler, D., T. Ernsting, M. Kerschensteiner, W. Bruck, and C. Stadelmann. 2006. ‘A new focal EAE model of cortical demyelination: multiple sclerosis-like lesions with rapid resolution of inflammation and extensive remyelination’, Brain, 129: 1972–83.

Metcalf, M., D. Xu, D. T. Okuda, L. Carvajal, R. Srinivasan, D. A. Kelley, P. Mukherjee, S. J. Nelson, D. B. Vigneron, and D. Pelletier. 2010. ‘High-resolution phased-array MRI of the human brain at 7 tesla: initial experience in multiple sclerosis patients’, J Neuroimaging, 20: 141–7.

Mike, A., B. I. Glanz, P. Hildenbrand, D. Meier, K. Bolden, M. Liguori, E. Dell’Oglio, B. C. Healy, R. Bakshi, and C. R. Guttmann. 2011. ‘Identification and clinical impact of multiple sclerosis cortical lesions as assessed by routine 3T MR imaging’, AJNR Am J Neuroradiol, 32: 515–21.

Mistry, N., R. Abdel-Fahim, O. Mougin, C. Tench, P. Gowland, and N. Evangelou. 2014. ‘Cortical lesion load correlates with diffuse injury of multiple sclerosis normal appearing white matter’, Mult Scler, 20: 227–33.

Moher, D., A. Liberati, J. Tetzlaff, D. G. Altman, and Prisma Group. 2009. ‘Preferred reporting items for systematic reviews and meta-analyses: the PRISMA statement’, PLoS Med, 6: e1000097.

Moll, N. M., A. M. Rietsch, A. J. Ransohoff, M. B. Cossoy, D. Huang, F. S. Eichler, B. D. Trapp, and R. M. Ransohoff. 2008. ‘Cortical demyelination in PML and MS: Similarities and differences’, Neurology, 70: 336–43.

Moraal, B., S. D. Roosendaal, P. J. Pouwels, H. Vrenken, R. A. van Schijndel, D. S. Meier, C. R. Guttmann, J. J. Geurts, and F. Barkhof. 2008. ‘Multi-contrast, isotropic, single-slab 3D MR imaging in multiple sclerosis’, Eur Radiol, 18: 2311–20.

Mougin, O., R. Abdel-Fahim, R. Dineen, A. Pitiot, N. Evangelou, and P. Gowland. 2016. ‘Imaging gray matter with concomitant null point imaging from the phase sensitive inversion recovery sequence’, Magn Reson Med, 76: 1512–16.

Nelson, F., S. Datta, N. Garcia, N. L. Rozario, F. Perez, G. Cutter, P. A. Narayana, and J. S. Wolinsky. 2011. ‘Intracortical lesions by 3T magnetic resonance imaging and correlation with cognitive impairment in multiple sclerosis’, Mult Scler, 17: 1122–9.

Nelson, F., A. H. Poonawalla, P. Hou, F. Huang, J. S. Wolinsky, and P. A. Narayana. 2007. ‘Improved identification of intracortical lesions in multiple sclerosis with phase-sensitive inversion recovery in combination with fast double inversion recovery MR imaging’, AJNR Am J Neuroradiol, 28: 1645–9.

Nelson, F., A. Poonawalla, P. Hou, J. S. Wolinsky, and P. A. Narayana. 2008. ‘3D MPRAGE improves classification of cortical lesions in multiple sclerosis’, Mult Scler, 14: 1214–9.

Nielsen, A. S., R. P. Kinkel, N. Madigan, E. Tinelli, T. Benner, and C. Mainero. 2013. ‘Contribution of cortical lesion subtypes at 7T MRI to physical and cognitive performance in MS’, Neurology, 81: 641–9.

Nielsen, A. S., R. P. Kinkel, E. Tinelli, T. Benner, J. Cohen-Adad, and C. Mainero. 2012. ‘Focal cortical lesion detection in multiple sclerosis: 3 Tesla DIR versus 7 Tesla FLASH-T2’, J Magn Reson Imaging, 35: 537–42.

Peterson, J. W., L. Bo, S. Mork, A. Chang, and B. D. Trapp. 2001. ‘Transected neurites, apoptotic neurons, and reduced inflammation in cortical multiple sclerosis lesions’, Ann Neurol, 50: 389–400.

Pisani, A. I., A. Scalfari, F. Crescenzo, C. Romualdi, and M. Calabrese. 2021. ‘A novel prognostic score to assess the risk of progression in relapsing-remitting multiple sclerosis patients’, Eur J Neurol.

Pitt, D., A. Boster, W. Pei, E. Wohleb, A. Jasne, C. R. Zachariah, K. Rammohan, M. V. Knopp, and P. Schmalbrock. 2010. ‘Imaging cortical lesions in multiple sclerosis with ultra-high-field magnetic resonance imaging’, Arch Neurol, 67: 812–8.

Pohmann, R., O. Speck, and K. Scheffler. 2016. ‘Signal-to-noise ratio and MR tissue parameters in human brain imaging at 3, 7, and 9.4 tesla using current receive coil arrays’, Magn Reson Med, 75: 801–9.

Popescu, B. F., J. E. Parisi, J. A. Cabrera-Gomez, K. Newell, R. N. Mandler, S. J. Pittock, V. A. Lennon, B. G. Weinshenker, and C. F. Lucchinetti. 2010. ‘Absence of cortical demyelination in neuromyelitis optica’, Neurology, 75: 2103–9.

Puthenparampil, M., D. Poggiali, F. Causin, G. Rolma, F. Rinaldi, P. Perini, and P. Gallo. 2016. ‘Cortical relapses in multiple sclerosis’, Mult Scler, 22: 1184–91.

Rovira, A., M. P. Wattjes, M. Tintore, C. Tur, T. A. Yousry, M. P. Sormani, N. De Stefano, M. Filippi, C. Auger, M. A. Rocca, F. Barkhof, F. Fazekas, L. Kappos, C. Polman, D. Miller, X. Montalban, and Magnims study group. 2015. ‘Evidence-based guidelines: MAGNIMS consensus guidelines on the use of MRI in multiple sclerosis-clinical implementation in the diagnostic process’, Nat Rev Neurol, 11: 471–82.

Saranathan, M., T. Tourdias, E. Bayram, P. Ghanouni, and B. K. Rutt. 2015. ‘Optimization of white-matter-nulled magnetization prepared rapid gradient echo (MP-RAGE) imaging’, Magn Reson Med, 73: 1786–94.

Saranathan, M., T. Tourdias, A. B. Kerr, J. D. Bernstein, G. A. Kerchner, M. H. Han, and B. K. Rutt. 2014. ‘Optimization of magnetization-prepared 3-dimensional fluid attenuated inversion recovery imaging for lesion detection at 7 T’, Invest Radiol, 49: 290–8.

Saranathan, M., P. W. Worters, D. W. Rettmann, B. Winegar, and J. Becker. 2017. ‘Physics for clinicians: Fluid-attenuated inversion recovery (FLAIR) and double inversion recovery (DIR) Imaging’, J Magn Reson Imaging, 46: 1590–600.

Sati, P. 2018. ‘Diagnosis of multiple sclerosis through the lens of ultra-high-field MRI’, J Magn Reson, 291: 101–09.

Scalfari, A., C. Romualdi, R. S. Nicholas, M. Mattoscio, R. Magliozzi, A. Morra, S. Monaco, P. A. Muraro, and M. Calabrese. 2018. ‘The cortical damage, early relapses, and onset of the progressive phase in multiple sclerosis’, Neurology, 90: e2107–e18.

Schmierer, K., H. G. Parkes, P. W. So, S. F. An, S. Brandner, R. J. Ordidge, T. A. Yousry, and D. H. Miller. 2010. ‘High field (9.4 Tesla) magnetic resonance imaging of cortical grey matter lesions in multiple sclerosis’, Brain, 133: 858–67.

Schmierer, K., J. R. Thavarajah, S. F. An, S. Brandner, D. H. Miller, and D. J. Tozer. 2010. ‘Effects of formalin fixation on magnetic resonance indices in multiple sclerosis cortical gray matter’, J Magn Reson Imaging, 32: 1054–60.

Schmierer, K., D. J. Tozer, F. Scaravilli, D. R. Altmann, G. J. Barker, P. S. Tofts, and D. H. Miller. 2007. ‘Quantitative magnetization transfer imaging in postmortem multiple sclerosis brain’, J Magn Reson Imaging, 26: 41–51.

Seewann, A., E. J. Kooi, S. D. Roosendaal, P. J. Pouwels, M. P. Wattjes, P. van der Valk, F. Barkhof, C. H. Polman, and J. J. Geurts. 2012. ‘Postmortem verification of MS cortical lesion detection with 3D DIR’, Neurology, 78: 302–8.

Seewann, A., H. Vrenken, E. J. Kooi, P. van der Valk, D. L. Knol, C. H. Polman, P. J. Pouwels, F. Barkhof, and J. J. Geurts. 2011. ‘Imaging the tip of the iceberg: visualization of cortical lesions in multiple sclerosis’, Mult Scler, 17: 1202–10.

Sepulcre, J., J. C. Masdeu, J. Goni, G. Arrondo, N. Velez de Mendizabal, B. Bejarano, and P. Villoslada. 2009. ‘Fatigue in multiple sclerosis is associated with the disruption of frontal and parietal pathways’, Mult Scler, 15: 337–44.

Sethi, V., N. Muhlert, M. Ron, X. Golay, C. A. Wheeler-Kingshott, D. H. Miller, D. T. Chard, and T. A. Yousry. 2013. ‘MS cortical lesions on DIR: not quite what they seem?’, PLoS One, 8: e78879.

Sethi, V., T. A. Yousry, N. Muhlert, M. Ron, X. Golay, C. Wheeler-Kingshott, D. H. Miller, and D. T. Chard. 2012. ‘Improved detection of cortical MS lesions with phase-sensitive inversion recovery MRI’, J Neurol Neurosurg Psychiatry, 83: 877–82.

Sinnecker, T., J. Dorr, C. F. Pfueller, L. Harms, K. Ruprecht, S. Jarius, W. Bruck, T. Niendorf, J. Wuerfel, and F. Paul. 2012b. ‘Distinct lesion morphology at 7-T MRI differentiates neuromyelitis optica from multiple sclerosis’, Neurology, 79: 708–14.

Sinnecker, T., P. Mittelstaedt, J. Dorr, C. F. Pfueller, L. Harms, T. Niendorf, F. Paul, and J. Wuerfel. 2012a. ‘Multiple sclerosis lesions and irreversible brain tissue damage: a comparative ultrahigh-field strength magnetic resonance imaging study’, Arch Neurol, 69: 739–45.

Springer, E., B. Dymerska, P. L. Cardoso, S. D. Robinson, C. Weisstanner, R. Wiest, B. Schmitt, and S. Trattnig. 2016. ‘Comparison of Routine Brain Imaging at 3 T and 7 T’, Invest Radiol, 51: 469–82.

Takai, Y., T. Misu, K. Kaneko, N. Chihara, K. Narikawa, S. Tsuchida, H. Nishida, T. Komori, M. Seki, T. Komatsu, K. Nakamagoe, T. Ikeda, M. Yoshida, T. Takahashi, H. Ono, S. Nishiyama, H. Kuroda, I. Nakashima, H. Suzuki, M. Bradl, H. Lassmann, K. Fujihara, M. Aoki, and M. O. G. antibody Disease Consortium Japan. 2020. ‘Myelin oligodendrocyte glycoprotein antibody-associated disease: an immunopathological study’, Brain, 143: 1431–46.

Tallantyre, E. C., P. S. Morgan, J. E. Dixon, A. Al-Radaideh, M. J. Brookes, P. G. Morris, and N. Evangelou. 2010. ‘3 Tesla and 7 Tesla MRI of multiple sclerosis cortical lesions’, J Magn Reson Imaging, 32: 971–7.

Thompson, A. J., B. L. Banwell, F. Barkhof, W. M. Carroll, T. Coetzee, G. Comi, J. Correale, F. Fazekas, M. Filippi, M. S. Freedman, K. Fujihara, S. L. Galetta, H. P. Hartung, L. Kappos, F. D. Lublin, R. A. Marrie, A. E. Miller, D. H. Miller, X. Montalban, E. M. Mowry, P. S. Sorensen, M. Tintore, A. L. Traboulsee, M. Trojano, B. M. J. Uitdehaag, S. Vukusic, E. Waubant, B. G. Weinshenker, S. C. Reingold, and J. A. Cohen. 2018. ‘Diagnosis of multiple sclerosis: 2017 revisions of the McDonald criteria’, Lancet Neurol, 17: 162–73.

Trapp, B. D., M. Vignos, J. Dudman, A. Chang, E. Fisher, S. M. Staugaitis, H. Battapady, S. Mork, D. Ontaneda, S. E. Jones, R. J. Fox, J. Chen, K. Nakamura, and R. A. Rudick. 2018. ‘Cortical neuronal densities and cerebral white matter demyelination in multiple sclerosis: a retrospective study’, Lancet Neurol, 17: 870–84.

Treaba, C. A., T. E. Granberg, M. P. Sormani, E. Herranz, R. A. Ouellette, C. Louapre, J. A. Sloane, R. P. Kinkel, and C. Mainero. 2019. ‘Longitudinal Characterization of Cortical Lesion Development and Evolution in Multiple Sclerosis with 7.0-T MRI’, Radiology, 291: 740–49.

Vercellino, M., F. Plano, B. Votta, R. Mutani, M. T. Giordana, and P. Cavalla. 2005. ‘Grey matter pathology in multiple sclerosis’, J Neuropathol Exp Neurol, 64: 1101–7.

Watkins, L. M., J. W. Neal, S. Loveless, I. Michailidou, V. Ramaglia, M. I. Rees, R. Reynolds, N. P. Robertson, B. P. Morgan, and O. W. Howell. 2016. ‘Complement is activated in progressive multiple sclerosis cortical grey matter lesions’, J Neuroinflammation, 13: 161.

Yao, B., S. Hametner, P. van Gelderen, H. Merkle, C. Chen, H. Lassmann, J. H. Duyn, and F. Bagnato. 2014. ‘7 Tesla magnetic resonance imaging to detect cortical pathology in multiple sclerosis’, PLoS One, 9: e108863.

Zurawski, J., S. Tauhid, R. Chu, F. Khalid, B. C. Healy, H. L. Weiner, and R. Bakshi. 2020. ‘7T MRI cerebral leptomeningeal enhancement is common in relapsing-remitting multiple sclerosis and is associated with cortical and thalamic lesions’, Mult Scler, 26: 177–87.

